# Cause-specific mortality attributable to summer heat in Italy, 2004–2023: a nationwide Bayesian spatiotemporal study

**DOI:** 10.64898/2026.10.02.26364563

**Authors:** Emiliano Ceccarelli, Giorgia Stoppa, Marta Blangiardo, Garyfallos Konstantinoudis, Nicola Vanacore, Giovanna Jona Lasinio, Giada Minelli

## Abstract

**Background:** Italy experiences a substantial heat-related mortality burden, but evidence on cause-specific mortality across its heterogeneous climatic settings remains limited. We aimed to quantify cause-specific mortality attributable to summer heat in Italy at national and ecological region levels and to map its distribution across municipalities.

**Methods:** We analysed daily deaths classified by underlying cause in eight ICD-10 groups among individuals aged 18–64 years and 65 years or older across 7,864 municipalities during June–August, 2004–2023. Mortality data were linked to area-weighted ERA5-Land daily maximum temperature. Nonlinear temperature–mortality associations were estimated with Bayesian hierarchical Poisson models, fitted separately for each cause, age group, and ecological region, with municipality-level attributable fractions.

**Findings:** High temperatures were associated with mortality from a broad range of causes, particularly among individuals aged 65 years or older. Among the study population, heat-attributable fractions were highest for nervous system diseases (14*·*93%, 95% credible inter-val 14*·*22–15*·*69) and mental and behavioural disorders (14*·*47%, 13*·*55–15*·*39). Circulatory diseases accounted for the largest absolute burden, with 85,524 heat-attributable deaths (82,762–88,271) across the summers of the 20 years considered.

**Interpretation:** The mortality burden associated with summer heat in Italy extended well beyond circulatory and respiratory diseases, highlighting the relevance of a broader range of health outcomes. These findings support heat–health strategies that account for cause-specific and geographical differences in heat-related risk, with greater consideration of neurological and mental health outcomes alongside established cardiorespiratory risks.

**Funding:** NextGenerationEU (CUP: B53C23002660006); Imperial College Research Fellowship; NIHR Imperial Biomedical Research Center (BRC: NIHR203323).

**Research in context:** *Evidence before this study:* We searched PubMed from database inception to 6 August 2026, combining exposure terms (“heat”, “high temperature”, or “ambient temperature”) with mortality terms (“mortality”, “cause-specific mortality”, or “cause of death”). Additional searches included terms related to attributable burden (“attributable fraction” or “attributable burden”) and geographical setting (“Italy” or “Europe”). We considered population-based observational studies examining mortality associated with high ambient temperatures, with particular attention to studies reporting cause-specific associations or attributable burden. Previous studies have consistently associated high ambient temperatures with increased mortality. Cause-specific analyses have shown that the effects of heat extend beyond cardiovascular and respiratory diseases to mental and behavioural, neurological, endocrine and metabolic, genitourinary, digestive, and external causes. However, much of the European evidence has been based on selected cities or relatively large administrative areas, and geographical variation has generally been characterised using administrative regions or broad climatic zones. Evidence on the fine-scale spatial distribution of cause-specific heat-attributable mortality across Italy remains limited, particularly when accounting for geographical differences in temperature–mortality associations.

*Added value of this study:* To our knowledge, this is the first nationwide Italian study to estimate cause-specific heat–mortality associations separately across ecological regions and to map the corresponding heat-attributable burden at the municipality level. Using daily mortality records from all Italian municipalities over the 20-year period 2004–2023, we estimated heat–mortality associations separately across ecological regions and age groups within a Bayesian modelling framework, propagating uncertainty from the estimated exposure–response associations to the attributable burden. We found consistent heat-related mortality risks across all major causes of death among older adults, with the strongest associations observed for mental and behavioural disorders and diseases of the nervous system.

*Implications of all the available evidence:* The available evidence supports considering a broader range of causes than cardiovascular and respiratory diseases alone in heat-health surveillance and prevention. Evidence of increased mortality for selected causes among adults aged 18–64 years also supports extending prevention beyond older populations. Accounting for heterogeneity across ecoregions reflects climatic variability across the country, provides a more accurate assessment of at-risk populations, and offers a framework applicable to other countries and regions characterised by similar climatic variability. These findings can support public health authorities in prioritising prevention measures and allocating resources where they are most needed under a warming climate.

## 1 Introduction

Exposure to high ambient temperatures is a major environmental risk factor for mortality, with a substantial heat-attributable burden documented worldwide and across European regions.^1^ In Europe, the summer of 2022 was associated with more than 60,000 heat-related deaths, ^2^ and more than half of heat-related deaths in 854 European cities during the summers of 2000–2025 were estimated to be attributable to human-induced warming. ^3^ Heat-related mortality is unevenly distributed across populations, with vulnerabilities varying by age and geography.^1,2^ Understanding these differential impacts is crucial for public health planning, targeted warnings, and ultimately protecting the vulnerable.

Studies on heat vulnerability have mainly focused on all-cause mortality and have reported differential impacts by demographic characteristics, including age and sex. ^4^ Cause-specific studies have more often focused on cardiovascular and respiratory mortality, ^5^ whereas only a handful have examined a broader range of causes of death. ^6,7^ A smaller body of evidence has also investigated the geographical variation of heat–mortality associations, reporting substantial differences across areas and suggesting a role for factors including green space, social deprivation, and urbanicity.^8^

In Italy, a substantial heat-related mortality burden occurs across markedly heterogeneous climatic and environmental settings, ^2^ from the Alpine and Apennine chains and the Padana Plain to Mediterranean coastal areas and the major islands. ^9^ Earlier Italian studies focused mainly on large cities and reported differences in heat vulnerability by age, pre-existing conditions, and socioeconomic position. ^10,11^ A nationwide study covering 2006–2015 reported cause-specific heat-attributable mortality by administrative region and urbanisation level^12^, and a recent small-area study described municipality-level variation in all-cause heat-related mortality among older adults. ^13^ However, cause-specific heat-related mortality in Italy has not yet been characterised across climatically coherent areas. Previous nationwide cause-specific estimates were based on administrative regions and urbanisation level, ^12^ while municipality-level analyses have so far focused on all-cause mortality. Stratification by ecoregions may therefore better capture geographical variation in temperature–mortality associations, while municipality-level data allow the corresponding attributable burden to be characterised at a finer spatial resolution.

In this nationwide study, we estimated mortality attributable to summer heat across eight cause-of-death groups among adults aged 18–64 years and 65 years or older in Italy during 2004–2023. We fitted separate Bayesian models on the five ecoregions, allowing heat–mortality associations to vary across climatically coherent areas, while accounting for lags, non-linearities, public holidays and spatiotemporal trends.

## 2 Methods

### 2.1 Study Area and Outcome

All 7,864 inland municipalities in Italy, as of 01-01-2025, were included in the study. The analyses were based on the Cause of Death Database maintained by the Statistical Service of the Italian National Institute of Health (ISS), which includes all diseases and conditions reported on death certificates, as recorded in the National Mortality Registry of the Italian National Institute of Statistics (ISTAT). The study period spanned from 01-01-2004 to 31-12-2023. For each death, the outcome was defined according to the underlying cause of death reported on the death certificate and coded using the International Classification of Diseases, 10th Revision (ICD-10). Individuals were classified into two age groups, 18–64 years and 65 years or older, obtained from ISTAT. Daily mortality counts were aggregated at the municipality level separately for each age group. Analyses were conducted for the eight following cause-specific mortality groups, according to ICD-10: endocrine, nutritional and metabolic diseases (E00–E90), mental, behavioural and neurodevelopmental disorders (F00–F99), diseases of the nervous system (G00–G99), diseases of the circulatory system (I00–I99), diseases of the respiratory system (J00–J99), diseases of the digestive system (K00–K93), diseases of the genitourinary system (N00–N99), and external causes (V01–Y98). The broad list of macro causes was chosen based on literature and to provide a comprehensive assessment of the heat–mortality association across the main diagnostic groups. ^6,7^ Analyses were restricted to the summer months, June to August.

#### 2.1.1 The ecoregions

To capture the marked environmental heterogeneity of Italy, analyses were stratified by ecoregions, defined using the ISTAT classification as broad, geographically coherent areas sharing similar ecological and biogeographical characteristics. ^9^ This stratification provided a parsimonious structure for estimating ecoregion-specific temperature–mortality associations and allowed for more flexible adjustment across similar environmental contexts. Figure S1 of the Supplementary Materials shows a map of Italy including the Ecoregions stratification adopted in this work: *Alpine Chain*, *Padana Plain*, *Apennine Chain*, *Tyrrhenian and the Islands*, *Apulia and the Adriatic Coast*.

#### 2.1.2 Exposure data

Temperature data were extracted from the ERA5-Land reanalysis dataset. ^14^ The dataset provides global coverage on a regular latitude–longitude grid at 0*·* 1*^◦^ ×* 0*·* 1*^◦^* resolution, approximately 9 km, and describes land and soil conditions up to a depth of 289 cm across four model layers. Hourly 2m Temperature values were extracted and summarised into daily maximum temperatures, to capture short-term exposure to high daytime temperatures. To account for both immediate and short-delayed effects of heat on mortality, exposure was represented by the average daily maximum temperature over lags 0–3.^15^ Finally, municipality-level values were obtained using area-weighted averages, as further explained in section S1.1 of the Supplementary Materials.

### 2.2 Statistical analysis

Models were fitted separately by ecoregion, age group, and cause of death. This allowed temperature–mortality associations, seasonal patterns, long-term trends,holiday effects, and residual spatial heterogeneity to differ across ecoregions. Daily municipality-level mortality counts were modelled using Bayesian hierarchical Poisson regression, with the annual age-specific municipal population from ISTAT included as an offset. The temperature–mortality association was modelled as a nonlinear function of the lagged temperature exposure using natural cubic splines, with knots at the 10th, 75th, and 90th percentiles of the ecoregion-specific temperature distribution. ^16,17^ Models included two linear covariates and two random effects: calendar year to account for long-term trends, an indicator for Sundays and bank holidays, a first-order random walk to capture residual within-summer seasonality, and a Besag–York–Mollié 2 (BYM2) municipality-level random effect to account for residual spatially structured and unstructured heterogeneity in baseline mortality.

For each ecoregion, age group, and cause of death, 1,000 posterior samples of the temperature–response association were obtained to propagate uncertainty through the subsequent estimation of the MMT and heat-attributable burden. The minimum mortality temperature (MMT) was identified, for each posterior sample, as the temperature corresponding to the minimum mortality risk between the 25th and 90th percentiles of the ecoregion-specific temperature distribution. ^18^ Exposure–response curves were then expressed relative to the corresponding sample-specific MMT. Posterior medians were used as point estimates and the 2*·*5th and 97*·*5th percentiles as 95% credible intervals.

Heat-attributable mortality was defined for temperatures above the sample-specific MMT. Temperature-specific attributable fractions were applied to the observed mortality counts to estimate heat-attributable deaths (ECH). We additionally calculated the fraction of deaths attributable to heat (AFH) and the cumulative heat-attributable excess mortality rate (ERH), expressed as deaths per 1,000 inhabitants over the 20-year study period. Posterior uncertainty in the exposure–response associations and MMTs was propagated throughout the estimation of all attributable-burden measures.

We assessed robustness using three alternative model specifications: (i) models without the BYM2 spatial component; (ii) models extending the exposure window to lags 0–5; (iii) and a specification based on fixed effects for each combination of day of the week, month, year, and municipality, equivalent to a time-stratified case-crossover design. Model fit was compared using the Widely Applicable Information Criterion (WAIC). Full model specifications, prior distributions, and mathematical details of the estimation of the exposure–response functions and attributable burden are provided in Section S1.2 of the Supplementary Materials.

### 2.3 Implementation

All analyses were conducted using R, version 4.4.2. ^19^ Municipality-level area weighted averages of the climatic and environmental data were computed using the `extract` function of the `terra` package. ^20^ Statistical models were implemented and fitted with `R-INLA`. ^21^ Models were fitted using the HPC TeraStat2, a high-performance computing infrastructure at the Department of Statistics of Sapienza University of Rome (see https://www.dss.uniroma1.it/it/HPCTerastat2).

## 3 Results

### 3.1 Descriptive analysis

The analysis included 1,823,566 deaths across the eight selected cause-of-death groups during the 20 summers: 162,332 among adults aged 18–64 years and 1,661,234 among those aged 65 years or older (Table 1). Circulatory diseases were by far the most frequent cause of death, particularly among individuals aged 65 years and older. In this age group, they were followed by respiratory diseases, endocrine, nutritional and metabolic diseases, and diseases of the nervous system. Among individuals aged 18–64 years, external causes stand out as the second most frequent category, followed by diseases of the digestive system. Excluding external causes, the crude numbers of deaths were generally more than ten times higher among individuals aged 65 years and older than among those aged 18–64 years. For more information about municipalities, population density and deaths by ecoregions see Tables S1–S3.

**Table 1:** Number of deaths by cause and age group in Italy, 2004–2023.

| Cause of death | 18–64 years | ≥65 years |
| --- | --- | --- |
| Endocrine, nutritional and metabolic diseases | 11,396 | 116,715 |
| Mental, behavioural, and neurodevelopmental disorders | 3,614 | 86,935 |
| Diseases of the nervous system | 9,883 | 115,416 |
| Diseases of the circulatory system | 64,356 | 935,929 |
| Diseases of the respiratory system | 8,369 | 181,232 |
| Diseases of the digestive system | 16,861 | 91,647 |
| Diseases of the genitourinary system | 2,637 | 53,114 |
| External causes | 45,216 | 80,246 |
| **Overall** | **162,332** | **1,661,234** |

Figure 1 shows the average summer daily maximum temperature over the study period by ecoregion and its spatial distribution, highlighting the marked climatic heterogeneity across Italy. The Alpine Chain was consistently the coolest ecoregion, whereas Apulia and the Adriatic Coast was the warmest. An increasing trend in summer temperatures over the study period was observed across all ecoregions.

**Figure 1:**
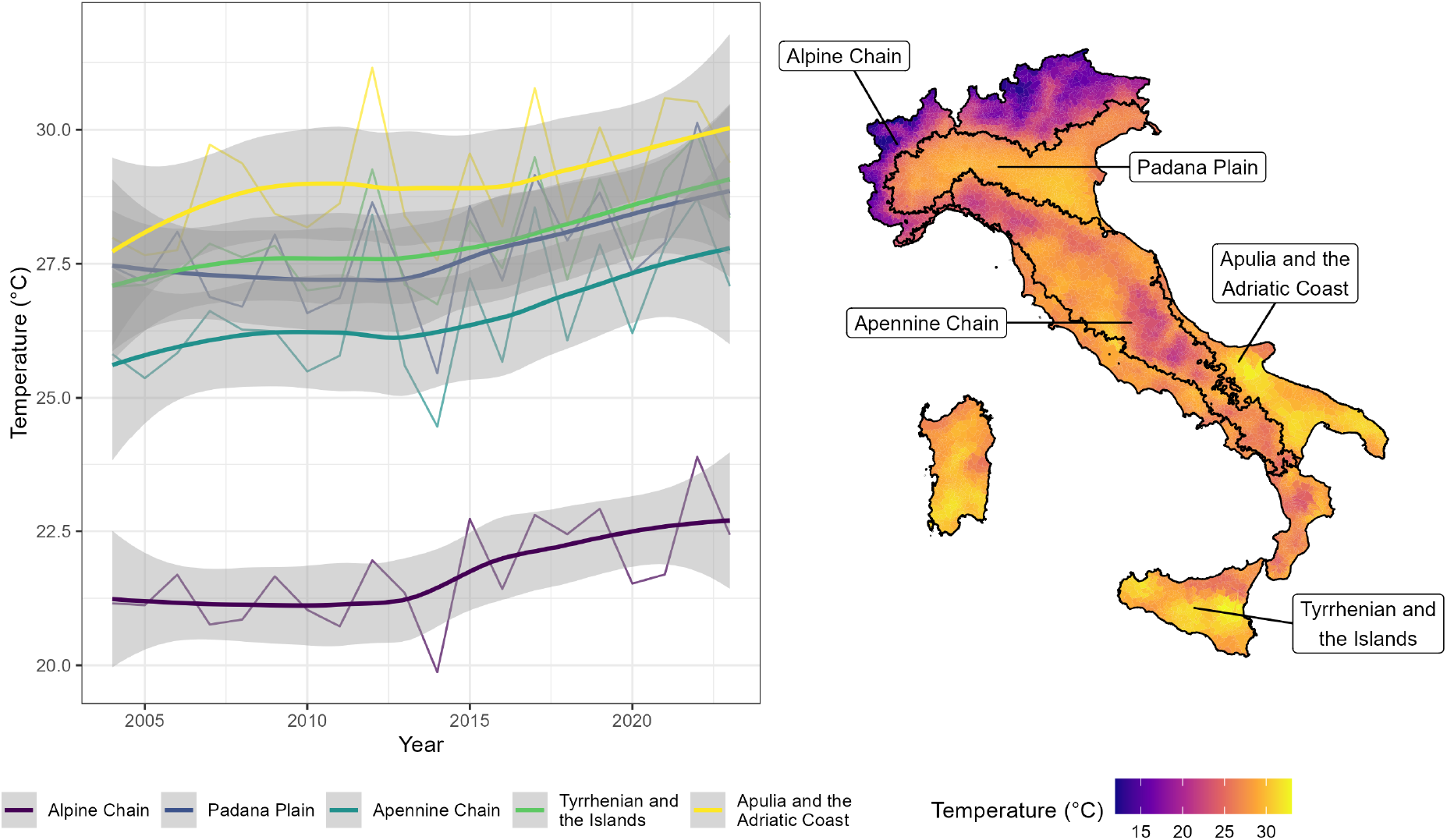
Left: annual average summer daily maximum temperature by ecoregion; the general trend is drawn using LOESS regression with a 95% confidence interval. Right: spatial distribution of average summer daily maximum temperature across municipalities over the study period, with ecoregion boundaries indicated.

### 3.2 Heat–mortality risk assessment and epidemiological metrics

Figure 2 shows the cause-specific relative risks at the 95th percentile of the ecoregion-specific daily maximum temperature distribution for both age groups. Among individuals aged 65 years and older, high temperatures were positively associated with mortality across causes and ecoregions, although the magnitude of the associations varied by cause and geographical area. The largest increases were generally observed for endocrine, nutritional and metabolic diseases, mental, behavioural and neurodevelopmental disorders, and diseases of the nervous system.

**Figure 2:**
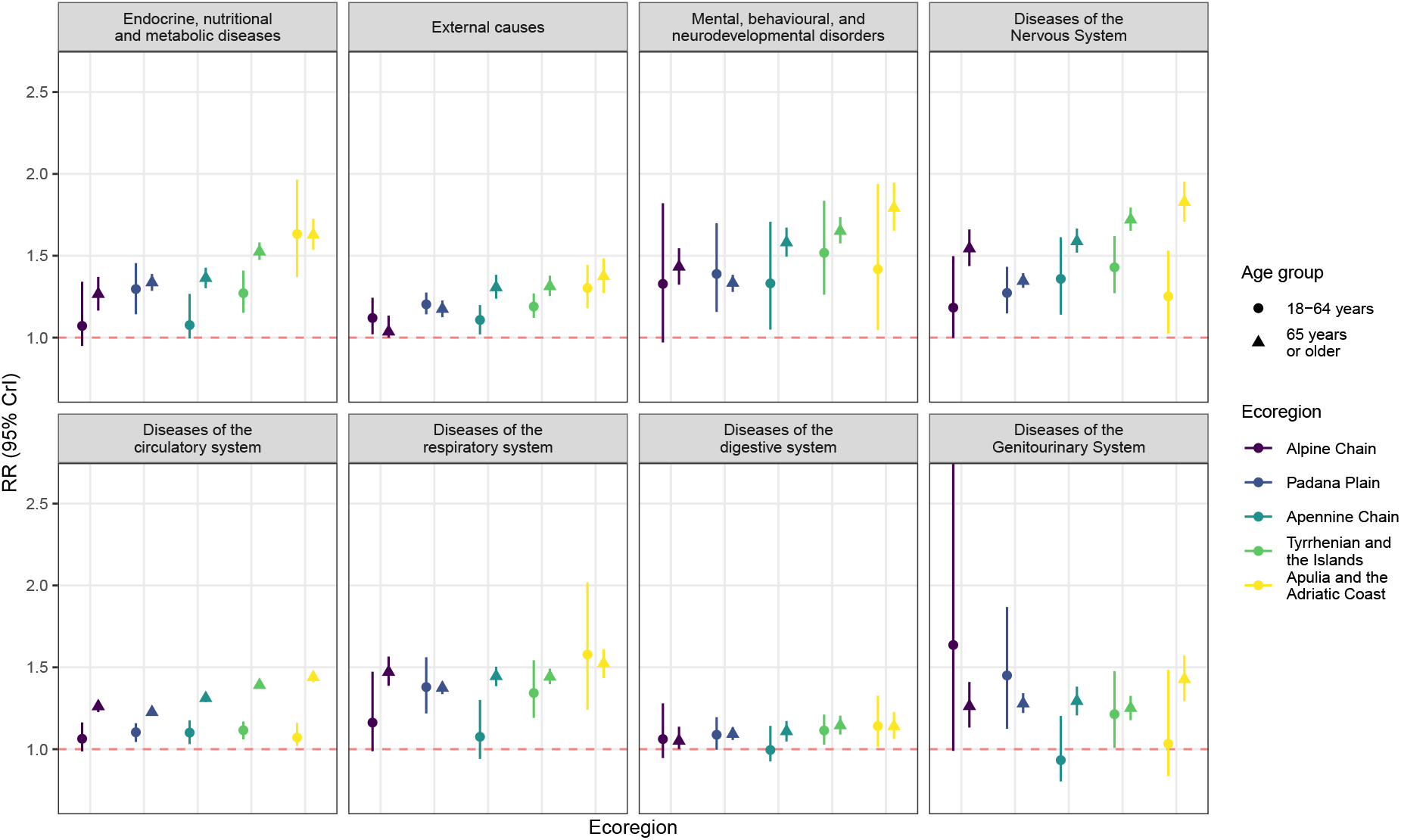
Cause-specific relative risk estimates at the 95th percentile of the ecoregion-specific daily maximum temperature distribution, by ecoregion and age group. Points represent posterior median estimates and vertical bars indicate 95% credible intervals.

A broadly similar geographical pattern was observed among individuals aged 18–64 years. For most causes, RR estimates were higher in the Tyrrhenian and the Islands and in Apulia and the Adriatic Coast, consistently with the patterns observed among individuals aged 65 years and older. However, estimates for the younger age group were substantially less precise, with wider credible intervals across several cause–ecoregion combinations.

The ecoregions are loosely capturing a north-to-south climatic gradient across Italy, and the observed exposure–response relationships showed a corresponding geographical pattern, with stronger heat-related increases in the southern and coastal ecoregions.

Figure 3 shows the exposure–response curves from the 1st to the 99th percentile of the ecoregion-specific temperature distribution for the eight causes of death considered among individuals aged 65 years and older. For most causes, the RR increased sharply in the upper tail of the temperature distribution, resulting in predominantly J-shaped relationships. The magnitude and shape of the associations nevertheless varied substantially across causes and ecoregions. External causes, diseases of the genitourinary system, diseases of the circulatory system, and diseases of the respiratory system showed clear increases at high temperatures, whereas digestive diseases showed the weakest heat-related association, with only a modest rise at the highest temperature percentiles. Full exposure–response curves for the 18–64 age group are reported in Figure S3 of the Supplementary Materials.

**Figure 3:**
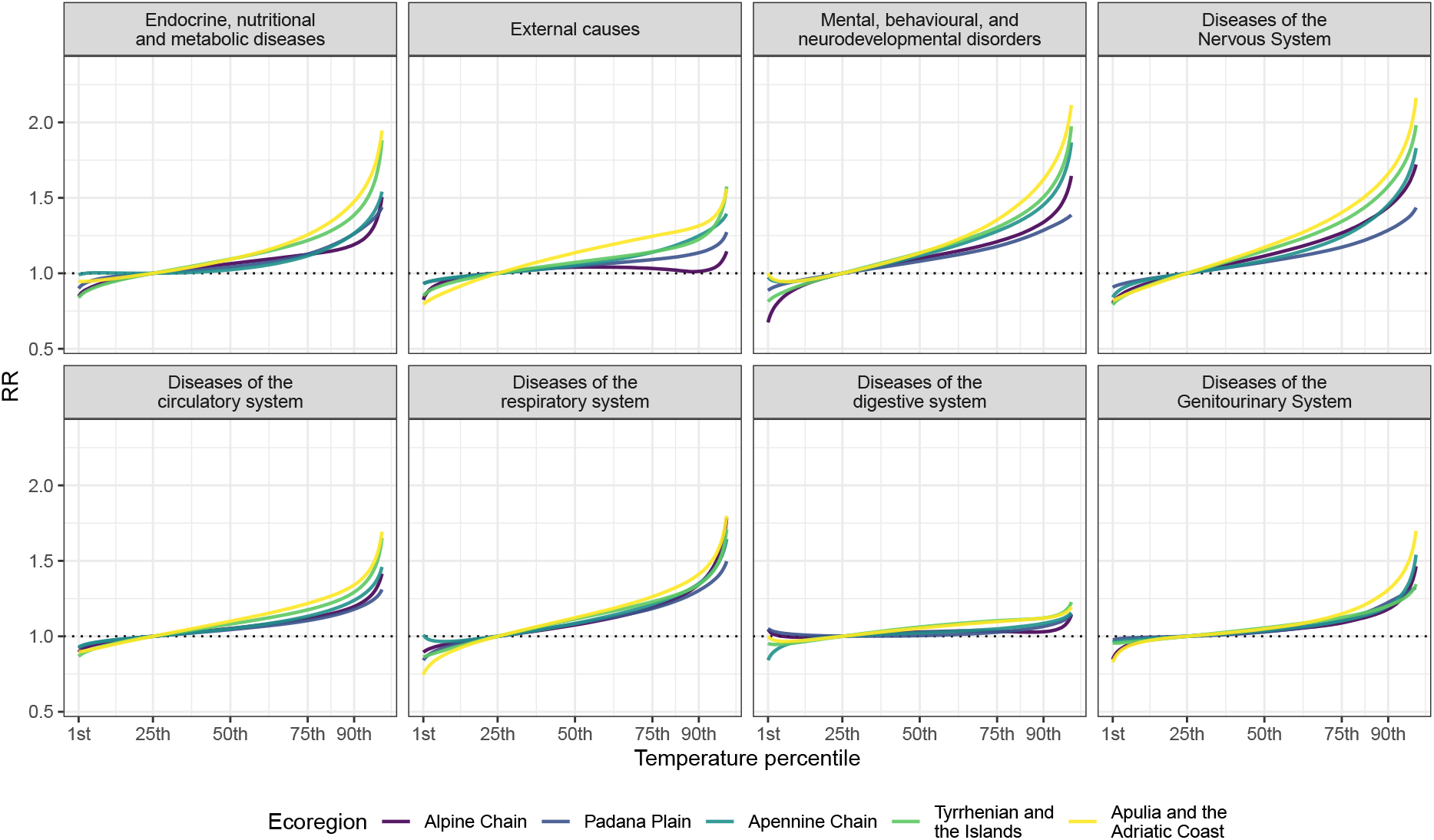
Cause-specific RR estimates for ecoregion-specific daily maximum temperature percentile exposure among individuals aged 65 years and older, by ecoregion. Curves span the 1st–99th percentiles of the temperature distribution.

The J-shaped curves also showed evident geographical heterogeneity across most causes. The Tyrrhenian and the Islands and Apulia and the Adriatic Coast generally showed the steepest increases at extreme temperatures. The Padana Plain exhibited flatter curves and lower maximum RRs, with relationships that were closer to linear for several causes, whereas the Alpine and Apennine Chains showed cause-specific patterns of intermediate magnitude. Detailed curves including the 95% credible intervals are reported in Figure S2 of the Supplementary Materials. MMT estimates were generally consistent across causes within the same ecoregion, with the lowest values in the Alpine Chain and the highest in Apulia and the Adriatic Coast (Figure S5). For several cause–age–ecoregion combinations, posterior MMTs lay at the lower bound of the search interval (the 25th percentile of the ecoregion-specific temperature distribution), suggesting that the minimum may lie at lower temperatures. Linear coefficients and random effects are reported in Section S2.1 of the Supplementary Materials.

Table 2 reports the heat-attributable fraction (AFH) and estimated count of heat-attributable deaths (ECH) across the eight causes of death and five ecoregions, combining the two age groups. At the national level, the largest AFHs were observed for diseases of the nervous system (14*·*93%, 95% credible interval 14*·*22–15*·*69), mental and behavioural disorders (14*·*47%, 13*·*55–15*·*39), and respiratory diseases (12*·*31%, 11*·*66–12*·*97). In contrast, circulatory diseases accounted for by far the largest absolute burden, with an estimated 85,524 heat-attributable deaths over the study period (82,762–88,271), followed by respiratory diseases (23,348, 22,113–24,590) and diseases of the nervous system (18,701, 17,821–19,665).

**Table 2:** Heat-attributable mortality by cause and ecoregion, all age groups combined.

|  | Alpine<br>Chain (1A) | Padana<br>Plain (1B) | Apennine<br>Chain (1C) | Tyrrhenian and<br>Islands (2B) | Apulia and<br>Adriatic Coast (2C) | Italy |
| --- | --- | --- | --- | --- | --- | --- |
| **Panel A. Heat-attributable fraction (AFH, %)** |  |  |  |  |  |  |
| Endocrine/metabolic | 9.47 (5.95–13.08) | 8.22 (6.97–9.55) | 8.52 (6.85–10.52) | 12.46 (11.25–13.74) | 15.37 (13.60–17.27) | 10.87 (10.16–11.63) |
| Mental/behavioural | 14.67 (10.98–18.08) | 10.30 (8.85–11.78) | 17.15 (14.91–19.52) | 17.15 (15.45–18.86) | 18.85 (15.60–21.52) | 14.47 (13.55–15.39) |
| Nervous system | 17.38 (14.38–20.66) | 10.11 (8.86–11.29) | 15.49 (13.67–17.38) | 17.19 (15.79–18.46) | 19.93 (18.03–22.03) | 14.93 (14.22–15.69) |
| Circulatory | 8.43 (7.22–9.61) | 6.19 (5.75–6.66) | 8.58 (7.92–9.28) | 9.72 (9.22–10.18) | 12.02 (11.16–12.84) | 8.55 (8.27–8.82) |
| Respiratory | 13.98 (11.33–16.63) | 10.50 (9.51–11.40) | 12.95 (11.29–14.52) | 12.51 (11.43–13.73) | 15.18 (13.19–16.99) | 12.31 (11.66–12.97) |
| Digestive | 2.63 (0.48–6.18) | 2.16 (1.34–3.37) | 3.37 (1.44–5.51) | 5.17 (3.59–6.72) | 5.25 (2.89–7.47) | 3.68 (2.93–4.53) |
| Genitourinary | 8.85 (4.33–14.43) | 7.12 (5.33–8.95) | 8.30 (5.37–11.14) | 7.64 (5.61–9.86) | 10.37 (7.06–13.78) | 7.96 (6.73–9.21) |
| External causes | 2.53 (0.70–5.42) | 6.66 (5.33–7.97) | 7.14 (5.48–8.90) | 8.52 (7.14–10.07) | 12.11 (9.77–14.21) | 7.48 (6.71–8.24) |
| **Panel B. Heat-attributable excess deaths (ECH)** |  |  |  |  |  |  |
| Endocrine/metabolic | 866 (544–1,195) | 2,737 (2,318–3,176) | 1,697 (1,365–2,096) | 6,384 (5,764–7,036) | 2,239 (1,981–2,517) | 13,927 (13,012–14,894) |
| Mental/behavioural | 1,506 (1,127–1,856) | 3,429 (2,945–3,919) | 2,456 (2,137–2,797) | 4,375 (3,942–4,810) | 1,353 (1,120–1,545) | 13,106 (12,273–13,934) |
| Nervous system | 2,127 (1,760–2,529) | 4,083 (3,579–4,562) | 3,269 (2,885–3,667) | 6,596 (6,059–7,082) | 2,629 (2,378–2,906) | 18,701 (17,821–19,665) |
| Circulatory | 7,896 (6,764–8,997) | 18,764 (17,411–20,158) | 14,442 (13,326–15,609) | 33,358 (31,635–34,921) | 11,105 (10,313–11,863) | 85,524 (82,762–88,271) |
| Respiratory | 2,506 (2,032–2,982) | 6,561 (5,947–7,128) | 4,162 (3,627–4,667) | 7,419 (6,780–8,140) | 2,692 (2,339–3,014) | 23,348 (22,113–24,590) |
| Digestive | 260 (48–611) | 786 (487–1,226) | 553 (237–903) | 1,838 (1,278–2,392) | 541 (298–770) | 3,996 (3,179–4,919) |
| Genitourinary | 418 (205–682) | 1,274 (953–1,602) | 749 (484–1,005) | 1,422 (1,045–1,836) | 568 (387–755) | 4,439 (3,753–5,134) |
| External causes | 330 (91–706) | 2,562 (2,050–3,065) | 1,472 (1,130–1,835) | 3,500 (2,933–4,140) | 1,484 (1,197–1,742) | 9,385 (8,421–10,332) |

Values are posterior medians (95% credible intervals). AFH, heat-attributable fraction; ECH, heat-attributable excess deaths. Estimates combine adults aged 18–64 years and those aged 65 years or older.

Consistently with the RR results, Apulia and the Adriatic Coast showed the highest AFH across all causes, with particularly large fractions for diseases of the nervous system (19*·*93%, 18*·*03–22*·*03), mental and behavioural disorders (18*·*85%, 15*·*60–21*·*52), and endocrine and metabolic diseases (15*·*37%, 13*·*60–17*·*27). The Tyrrhenian and the Islands also showed comparatively high attributable fractions and accounted for the largest number of heat-attributable deaths for every cause considered. In particular, circulatory diseases contributed an estimated 33,358 attributable deaths in this ecoregion alone (31,635–34,921).

Age-specific and ecoregion-specific estimates for the three epidemiological metrics are reported in Supplementary Tables S4–S9. Overall, heat-attributable burdens were substantially greater among individuals aged 65 years and older, both in relative and absolute terms. Among individuals aged 18–64 years, external causes had an attributable fraction of 6*·*57% (5*·*52–7*·*67) and accounted for the largest absolute burden, with an estimated 2,971 heat-attributable deaths (2,494–3,469).

To further characterise the geographical distribution of the heat-attributable burden among older adults, Figure 4 presents the posterior median municipality-level AFH estimates for the 65+ age group. The maps show broadly consistent spatial patterns across the different causes of death. These patterns reflect the combined effects of the ecoregion-specific exposure–response functions and the local distributions of temperature and mortality. The spatial distribution also suggested differences between inland and coastal municipalities.

**Figure 4:**
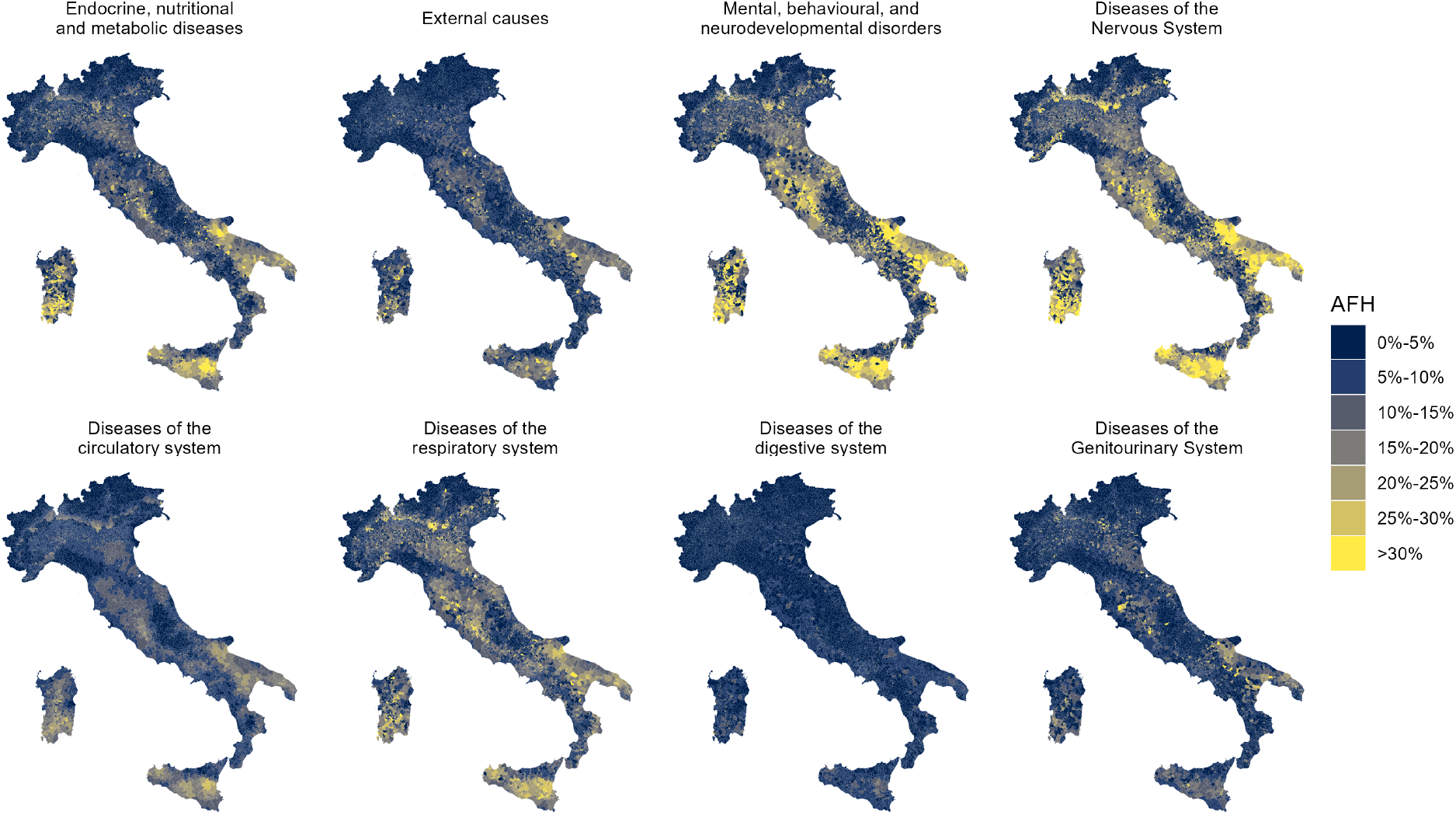
Heat-attributable fraction (AFH) by cause of death among individuals aged 65 years and older, by municipality. Values represent posterior median estimates.

Municipality-level results for the 18–64 age group are reported in Figure S8 of the Supplementary Materials and are characterised by substantially greater uncertainty in the estimated heat–mortality associations.

### 3.3 Sensitivity model results

Sensitivity analyses were conducted for the 65+ age group, as this group accounted for the largest number of deaths and showed the most robust and consistent heat–mortality associations in the main analysis. The results, shown in Figures S9, S10, and S11 of the Supplementary Materials, were largely consistent with those of the main model across all ecoregions and causes of death. In particular, extending the exposure window to lag 0–5 produced virtually identical exposure–response relationships, whereas omitting the spatial component or adopting a case-crossover-like temporal specification resulted in only modest differences, while preserving the overall shape of the associations. The corresponding municipality-level AFH maps (Figures S12 – S14 of the Supplementary Materials) showed consistent geographical patterns across sensitivity analyses, with modest differences in the magnitude of the attributable fractions, in line with the observed differences in the sensitivity models estimated exposure–response relationships. Model comparison based on the WAIC consistently favoured the main model over both the no-BYM2 and case-crossover specifications (Tables S10 and S11 of the Supplementary Materials).

## 4 Discussion

In this nationwide study covering 20 years, five Italian ecoregions and 7,864 municipalities, high temperatures were associated with mortality from a broad range of causes, particularly among individuals aged 65 years or older. The largest heat-attributable fractions were observed for diseases of the nervous system and mental and behavioural disorders. Positive associations were also observed among individuals aged 18–64 years for selected causes, although estimates were less precise. Heat-attributable fractions showed substantial spatial variation across the study region, with broadly consistent geographic patterns across diagnostic groups.

Our findings are consistent with previous evidence showing cause-specific differences in heat-related mortality. A previous nationwide Italian study conducted over 2006–15 reported a similar cause-specific pattern, with the largest heat-attributable fractions for neurological diseases (3*·*6%, 95% CI 1*·*9–4*·*9), mental disorders (3*·*1%, 1*·*7–4*·*4), respiratory diseases (2*·*4%, 1*·*4–3*·*4), and endocrine and metabolic diseases (2*·*3%, 1*·*1–3*·*4).^12^ Similar associations have also been reported in England and Wales and the USA for mental, neurological, endocrine, genitourinary, digestive, and external causes, although the magnitude of the estimated burden varied across populations and analytical designs. ^6,7^ In the USA, substantially lower fractions of mortality attributable to high temperatures were reported, for circulatory diseases (0*·*2%, 95% CI 0*·*1–0*·*3) and external causes (6*·*0%, 2*·*1–9*·*8). ^7^ In England and Wales, were estimated 8,005 heat-attributable deaths from circulatory diseases, 5,841 from respiratory diseases, and 1,118 from nervous system diseases over 14 summers, corresponding to approximately 572, 417, and 80 deaths per summer, respectively, compared with approximately 4,276, 1,167, and 935 deaths per summer in our study.^6^ Differences in the magnitude of these estimates should, however, be interpreted in light of differences in population size, study period, exposure metrics, definitions of heat, and the methods used to quantify attributable mortality.

Temperature–mortality associations differed across ecoregions, with stronger increases at the upper end of the local temperature distribution observed in the southern and coastal ecoregions. These comparisons should, however, be interpreted cautiously because the same temperature percentile corresponds to different absolute temperatures across ecoregions. A nationwide Italian small-area study estimated municipality-level spatial variation in heat-related mortality, covering the period 2011–2023 and focusing on all-cause mortality among older adults, highlighting similar substantial small-area differences in heat-related mortality across Italy and identifying socioeconomic deprivation as a key determinant of vulnerability.^13^ The observed spatial discrepancies may also reflect population characteristics, housing, access to cooling, occupational exposure, and other environmental or behavioural factors.

For circulatory and respiratory diseases, several physiological mechanisms may contribute to the observed heat-related burden. Heat can impose additional cardiovascular strain through changes in cardiac workload, blood viscosity, coagulation, and cerebral perfusion, whereas dehydration and the increased ventilatory effort required for heat dissipation might aggravate chronic respiratory disease. ^5,22^ Endocrine diseases also showed a substantial attributable burden. These findings are compatible with additional metabolic and renal stress arising from dehydration, electrolyte imbalance, and impaired physiological adaptation during periods of high temperature. ^23^ For external causes, the observed associations likely reflect both physiological and behavioural mechanisms. Heat exposure can impair cognitive performance, attention and decision-making through dehydration and heat stress, while also increasing fatigue and irritability. At the same time, warmer weather encourages outdoor activities and mobility, increasing exposure to hazards such as traffic accidents, falls and drowning. ^24^ The association observed for genitourinary diseases may partly reflect renal effects of heat-related fluid depletion, which can reduce renal blood flow and increase the risk of acute kidney injury.^25^ Digestive diseases showed the weakest association with high temperatures; however, the limited cause-specific epidemiological evidence precludes a firm interpretation of this finding.

These physiological mechanisms provide a plausible basis for the observed associations, but the magnitude of the burden estimated for each broad cause-of-death group also depends on the specific conditions contributing to that ICD-10 group.

Studying broad ICD-10 chapters requires caution when interpreting the results, as they include conditions with markedly different aetiologies. For example, in the study period, endocrine diseases were largely driven by diabetes mellitus (ICD-10 E10–E14), accounting for 78*·*6% of deaths among individuals aged 65 years or older and 61*·*3% among those aged 18–64 years, while obesity (E66) represented a further 16*·*4% of endocrine deaths in the younger age group. The mental and behavioural disorders chapter was highly homogeneous among older adults, with dementias (F00–F03) accounting for 94% of deaths, whereas deaths among individuals aged 18–64 years were much more heterogeneous, with alcohol- and substance-related disorders (F10 and F19) together representing 40*·*6% of cases. Diseases of the nervous system also showed substantial heterogeneity: Alzheimer’s disease accounted for 43*·*2% of deaths and Parkinson’s disease for 26*·*0% among older adults, whereas no single diagnosis predominated in the younger age group, where spinal muscular atrophy (G12) and multiple sclerosis (G35) were the most frequent causes. The interpretation of mental and nervous system diseases also requires consideration of the ICD-10 classification itself. Dementia is predominantly classified within mental and behavioural disorders (F00–F03), whereas Alzheimer’s disease (G30) is included among diseases of the nervous system. Thus, although these conditions are clinically and epidemiologically closely related, their associated deaths contribute to two different broad cause-of-death groups in our analysis. This distinction should be considered when interpreting the particularly high heat-related burden observed for both groups.

When interpreted alongside existing evidence, these findings may contribute to refining heat–health policies and support the implementation of heat–health action plans (HHAPs). Consistent with WHO guidance, ^26^ HHAPs encompass interconnected components including governance, heat–health early warning systems, identification of high-risk populations, communication, health-system resilience, reduction of heat exposure, heat–health surveillance, and monitoring, evaluation, and learning. Within this framework, preparedness can rely on continuous monitoring, predefined actions, and progressively intensified interventions as heat-related risks increase, complemented by longer-term, cross-sectoral measures aimed at reducing population exposure to heat and addressing environmental determinants of vulnerability. The observed cause-specific patterns support greater consideration of neurological and mental health outcomes, while the wider literature points to the potential relevance of cognitive impairment, multimorbidity, and treatments affecting thermoregulation or fluid balance. Preventive resources should be proportionate to estimated risks and account for barriers that may limit access to information, health care, and effective protection from heat.

The nationwide coverage and long observation period allowed us to investigate eight broad cause-of-death groups across two age groups while retaining sufficient information to characterise geographical heterogeneity. Rather than assuming a single national temperature–mortality relationship, we estimated separate nonlinear exposure–response functions across five ecoregions and applied them at the municipality level to quantify the heat-attributable burden at the highest spatial resolution available in the country. Finally, the Bayesian framework allowed uncertainty to be propagated from the dose–response estimates to the heat-attributable fraction. The distribution of deaths across the causes considered in this study was broadly consistent with the overall mortality profile reported for the European Union, ^27^ supporting the broader relevance of this cause-specific assessment.

Various limitations should also be considered. First, this ecological study used municipality-level temperature, which cannot represent individual exposure. Furthermore, temperature measurement error remains a potential source of bias. In particular, in complex terrain such as mountainous areas, grid-cell averaging and elevation mismatches between ERA5-Land pixels and urban centres may lead to misclassification of exposure. The broad age categories considered may have masked heterogeneity within the population, particularly among those aged 65 years or older. Modelling temporal changes using a single linear yearly term may not have captured non-linear trends in cause-specific mortality over the 20-year study period, including changes in disease incidence, diagnosis, treatment, and coding practices. We considered only the underlying cause of death reported on death certificates, and not other contributing conditions; this might fail to fully capture the contribution of conditions frequently reported as multiple causes. This limitation may be particularly relevant for neurodegenerative diseases, which are often recorded as contributing rather than underlying causes. In Italy, the ratios of multiple to underlying causes of death have been estimated at 1*·*7 for Alzheimer’s disease, 2*·*3 for other dementias, and 2*·*8 for Parkinson’s disease. ^28^ Furthermore, death certificates have shown limited sensitivity in identifying dementia and parkinsonism when compared with clinical information. ^29,30^ Consequently, the mortality burden associated with these conditions may be underestimated or assigned to other underlying cause-of-death categories.

## 5 Conclusions

Heat-related mortality extended well beyond circulatory and respiratory diseases, with particularly large attributable fractions for diseases of the nervous system and mental and behavioural disorders among older adults, while the magnitude of the associations varied across ecoregions. These findings highlight the importance of considering both the underlying cause of death and geographical variation when characterising the mortality burden associated with high temperatures. By identifying cause-specific and geographical differences in heat-related mortality, these results may also contribute to a more detailed characterisation of populations particularly susceptible to high temperatures.

## Contributors

E.C., G.J.L. and G.M. conceptualised the study. M.B. and G.K. refined the study design and analytical approach. E.C. conducted the analysis. G.S., G.M., G.J.L., M.B. and G.K. supervised. E.C. and G.S. wrote the initial draft. G.J.L., G.K., M.B., and N.V. provided valuable insights into the interpretation of the results. All authors contributed to reviewing and editing. G.M. was responsible for funding acquisition. G.M. and E.C. were responsible for the decision to submit and subsequent revisions.

## Acknowledgements and Funding

E.C. was supported by the European Union – NextGenerationEU, under the PNRR project “C_PA - DM118 P.A. Pubblica Amministrazione: Metodologie statistiche per il supporto alle decisioni in contesto sanitario pubblico: stima dell’impatto degli eventi climatici estremi sulla salute della popolazione generale e la costruzione di modelli di allerta rapida", CUP: B53C23002660006, Missione 4, Componente 1 (Missione I.4.1, PNRR Scholarships for Public Administration). G.K is supported by an Imperial College Research Fellowship. M.B acknowledges infrastructure support for the Department of Epidemiology and Biostatistics provided by the NIHR Imperial Biomedical Research Centre (BRC).

## Conflicts of interest

The authors declare no conflicts of interest.

## Data sharing

ERA5-Land data can be obtained from the Copernicus Climate Data Store at https://cds.climate.copernicus.eu/datasets/reanalysis-era5-land?tab=overview. Mortality data are available from the Istituto Superiore di Sanità, but restrictions apply; these data were used under licence for the current study and are therefore not publicly available. Data are, however, available from the authors upon reasonable request and with permission of Istituto Superiore di Sanità. Requests for access to the mortality dataset should be directed at. The code to replicate the analyses is available on GitHub at https://github.com/emilianoceccarelli/Cause_Specific_Heat_Italy.

## Supplementary Materials

### S1 Supplementary Methods

#### S1.1 Area-weighted averages

Given {*C*_1_*,…, C_K_*}, the ERA5-Land grid cells that fall within Italy, the temperature for municipality *m* on day *ℓ* and year *t* is given by:

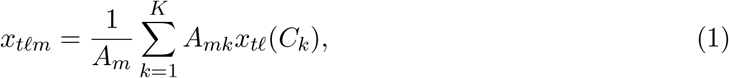

where *A_m_* is the total surface area of municipality *m*, for *m* = 1*,…,* 7,864, *A_mk_* is the area of the intersection between grid cell *C_k_* and municipality *m* and *x_tℓ_*(*C_k_*) is the recorded temperature in day *ℓ*, year *t* in the ERA5-Land cell *C_k_*. For grid cells that do not intersect municipality *m*, *A_mk_* = 0. The total surface area of the municipality can therefore be expressed as 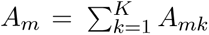.

#### S1.2 Main model

Models were estimated separately for each ecoregion, age group, and cause of death. For simplicity, the age-group and cause-specific indices are omitted from the notation. Let *Y_tℓm_* be the count of deaths occurring on day *ℓ* = 1*,…,* 92, year *t* = 1*,…,* 20, and municipality *m* = 1*,…,* 7, 864. For each ecoregion *e* = “1A”, “1B”, “1C”, “2B”, “2C”, age class *g* =“18–64 years”, “65+ years”, and each cause of death *c*, we specified the following Bayesian hierarchical Poisson model:

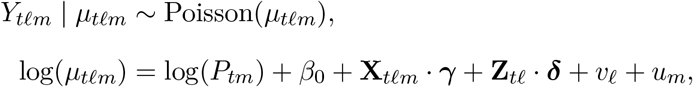

where *P_tm_* is the population of municipality *m* in year *t* for the corresponding age group; *β*_0_ is the global intercept; **X***_tℓm_ · **γ*** is a nonlinear function of the average daily maximum temperatures across lags 0–3^15^, represented by a matrix of basis functions, **X***_tℓm_*, and its coefficients, ***γ***; **Z***_tℓ_* is a matrix of linear covariates included in the model, presented in the following section, while ***δ*** are the corresponding regression coefficients; lastly, *v_ℓ_* and *u_m_* are random effects accounting for unmeasured temporal and spatial confounding, respectively. Specifically, *v_ℓ_* models the residual seasonal pattern, whereas *u_m_* follows a Besag–York–Mollié 2 (BYM2) specification, accounting for both spatially structured and unstructured heterogeneity in baseline mortality across municipalities.

The nonlinear relationship between temperature and mortality was modelled using natural cubic splines^16,17^, with knots at the 10th, 75th, and 90th percentiles of the ecoregion-specific temperature distribution and no intercept, so as to capture within-ecoregion climatic heterogeneity and focus on the upper tail.

The matrix **Z***_tℓ_* contains two linear covariates: the calendar year *t*, to account for long-term temporal trends, and a binary variable equal to 1 if the *ℓ*-th day fell on a bank holiday or a Sunday, and 0 otherwise.

The seasonality term *v_ℓ_* was modelled as a first-order random walk,

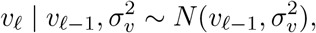

where 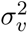 is the variance of the process.

Spatial dependence was modelled using the Besag-York-Mollié 2 (BYM2) model, which decomposes the municipality-specific effect into spatially structured and unstructured components,

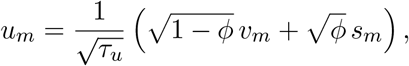

where *v_m_ ∼ N* (0, 1) represents the unstructured random effect, *s_m_* is an intrinsic conditional autoregressive (ICAR) spatial effect defined according to the municipality adjacency structure, *τ_u_* is the overall precision parameter, and *ϕ ∈* [0, 1] controls the proportion of total variance explained by the spatially structured component.

##### S1.2.1 Prior and Hyperprior specification

Penalised complexity (PC) priors were assigned to the precision parameters of the random effects. For the first-order random walk term modelling within-season seasonality, we used a PC prior on 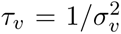 with *P* (*σ_v_ >* 0.3) = 0.01. The overall precision parameter *τ_u_* of the BYM2 was assigned a PC prior such that

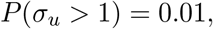

where 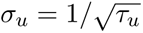, while the mixing parameter *ϕ*, controlling the proportion of spatially structured variance, was assigned the default PC prior

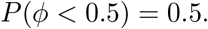

Independent Gaussian priors were assigned to the fixed effects ***γ***, ***δ*** *∼ N* (0, 1000), while the intercept was assigned the default improper flat prior *β*_0_ *∼ N* (0*, ∞*).

#### S1.3 Temperature–mortality association

For each ecoregion, we defined a temperature distribution **x***_e_* spanning from the minimum to the maximum observed temperature, with increments of 0.1 percentiles. The corresponding basis matrix, **X***_e_ ∈* ℝ^1001×4^, was initially centred at the ecoregion-specific 75th percentile of the temperature distribution, 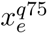, by subtracting the basis vector evaluated at 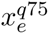:

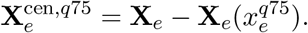

We drew 1,000 samples of the temperature–response coefficients, 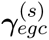, from the model’s posterior distribution and calculated the preliminary temperature-related relative risk for each ecoregion *e*, age class *g*, cause of death *c*, and posterior sample *s*, after accounting for confounding, as follows:

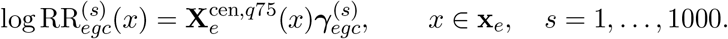

The resulting sample-specific exposure–response curves were used to identify the sample-specific minimum mortality temperature, 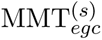, defined as the temperature corresponding to the minimum log RR within the 25th–90th percentiles of the ecoregion-specific temperature distribution, consistently with previous studies^17^.

Finally, the basis matrix **X***_e_* was re-centred separately by subtracting the basis vector evaluated at the corresponding 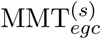, yielding the final centred basis matrices. The posterior logRR curves were then recomputed using the re-centred basis and exponentiated, so that each exposure–response curve was expressed relative to its corresponding 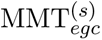.

At each temperature value, the posterior median of the 1,000 RR estimates was reported as the point estimate, while the 2.5th and 97.5th percentiles were used to construct the 95% credible interval.

Uncertainty was propagated throughout the attributable fraction analysis by retaining the full set of 1,000 posterior RR curves for each ecoregion, age class, and cause of death.

##### S1.3.1 Excess mortality rates attributable to heat

We computed two epidemiological metrics at the municipality level: the excess mortality rates attributable to heat (ERH) and the Fraction of deaths attributable to heat (AFH). To do so, we first set the 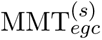 as threshold temperature and computed the attributable fraction (AF) for each ecoregion, age class and cause, and for the temperature values *x*:

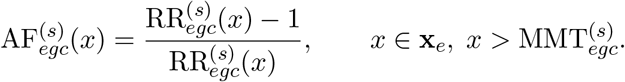

Let *e*(*m*) denote the ecoregion to which municipality *m* belongs. We obtained the excess death counts attributable to heat (ECH) at the municipality level by multiplying the AF by the death counts that occurred at temperature *x*, *Y_mc_*(*x*):

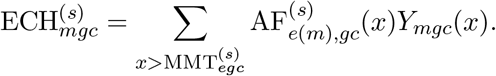

Finally, we obtained the standardised metrics by dividing the ECH by the population or the total number of observed deaths:

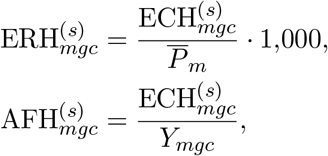

where *P̅_mg_* is the average population of the *g*th age class of municipality *m* over the 20-year study period, representing the cumulative heat-attributable deaths per 1,000 average residents over 2004–2023. From the sample-specific estimates, we used the posterior median as point estimates and computed the 95% credible intervals.

##### S1.3.2 Sensitivity analysis

To assess the robustness of the estimated heat–mortality association, we compared the main results with those obtained under three alternative model specifications: (i) removing the BYM2 component, to evaluate the impact of accounting for spatial confounding; (ii) extending the exposure window from lags 0–3 to lags 0–5, to investigate a potentially longer-lasting heat effect; and (iii) replacing the year fixed effect and the temporal and spatial random effects with a set of fixed effects for each unique combination of day-of-week, month, year, and municipality — an approach equivalent to a time-stratified case-crossover design, allowing seasonality and long-term trends to vary across spatial units. The complete mathematical formulation of each sensitivity model is reported Section S1.4. Models were compared both in terms of the estimated heat–mortality associations and their overall goodness-of-fit, assessed using the Widely Applicable Information Criterion (WAIC). WAIC provides an estimate of out-of-sample predictive accuracy while accounting for model complexity and is defined as

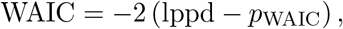

#### where the log pointwise predictive density (lppd) is

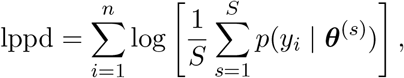

and the effective number of parameters is estimated as

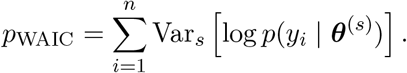

Here, *i* indexes the observations, *s* = 1*,…, S* indexes posterior samples, and ***θ***^(^*^s^*^)^ denotes the corresponding posterior draw of the model parameters. Lower WAIC values indicate better expected predictive performance, with comparisons being made between models fitted to the same data. For ease of comparison, we report ΔWAIC = WAIC_main_ *−* WAIC_sensitivity_; therefore, negative values indicate better predictive performance of the main model.

#### S1.4 Sensitivity analysis models

##### S1.4.1 Sensitivity analysis model 1

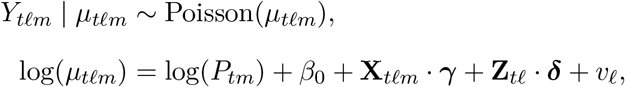

##### S1.4.2 Sensitivity analysis model 2

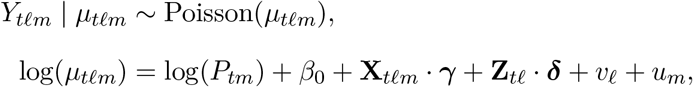

**X***_tℓm_ ·* ***γ*** is, in this case, a nonlinear function of the average daily maximum temperatures across lags 0–5.

##### S1.4.3 Sensitivity analysis model 3

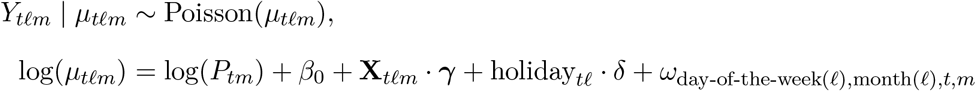

where *ω*_day*−*of*−*the*−*week(*t*),month(*ℓ*)*,t,m*_ denotes a fixed effect indexing each unique combination of day-of-the-week and month of the *ℓ*-th day, calendar year *t*, and municipality *m*.

### S2 Supplementary results

#### S2.1 Linear coefficients and random effects

Overall, no consistent association with holidays was observed, with most estimates close to the null and wide credible intervals. Conversely, a generally positive linear yearly trend was estimated for most causes, especially among individuals aged 65 years and older, suggesting increasing mortality over the study period. Exceptions were circulatory diseases, for which a weak negative trend was estimated in most ecoregions, and digestive diseases, which showed little evidence of temporal changes.

Figures S6 and S7 show the main model’s random effect results: respectively, the estimated seasonality effect — in terms of RRs — and the posterior probability that the municipality-level residual spatial RR is above one. The seasonal component showed no clear effect in the majority of the causes and both age classes, except for cardiovascular and respiratory in the 65+ age group, where a mild decreasing trend is estimated.

The spatial component showed a not-negligible residual spatial heterogeneity. Areas with higher posterior probability of residual risk were scattered across the country, with broadly similar spatial patterns for the different causes.

### S3 Supplementary figures

**Figure S1:**
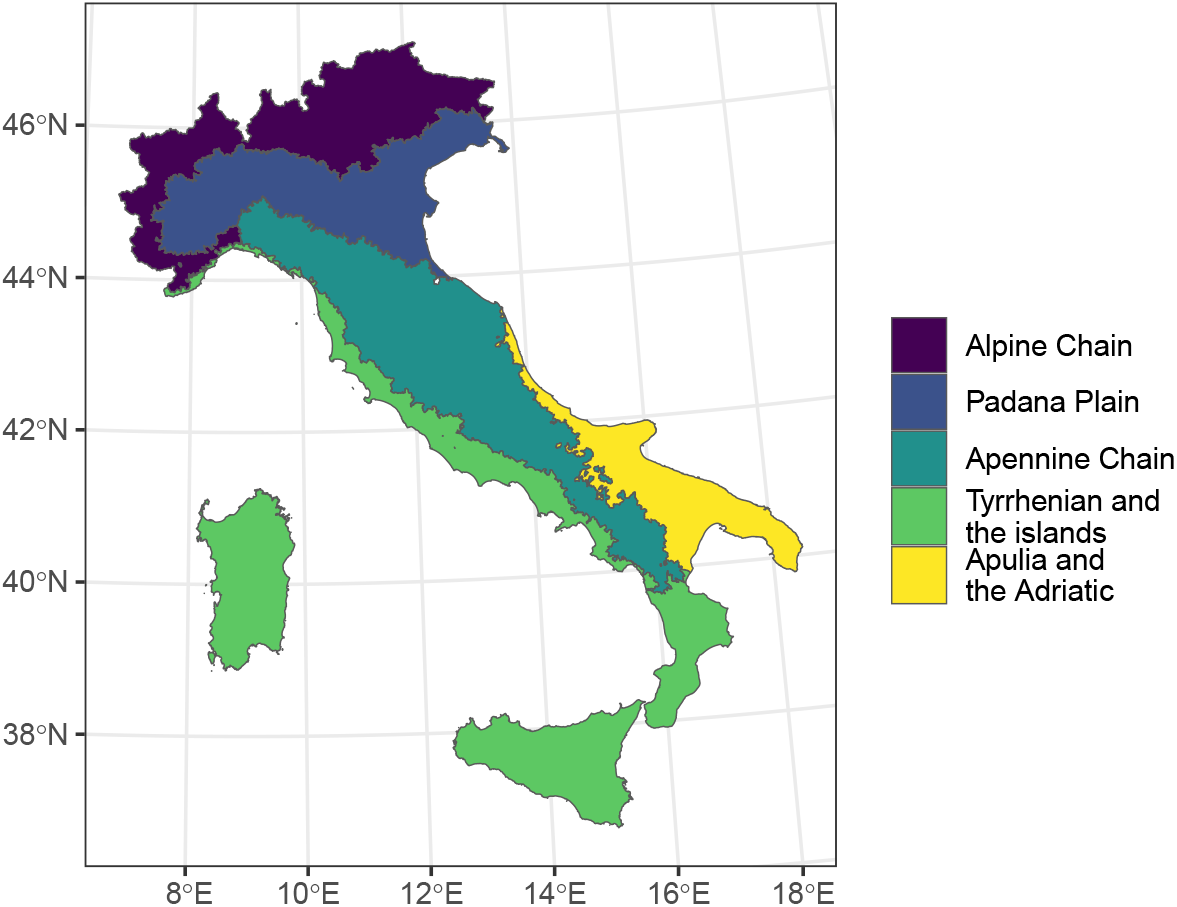
Map of Italy by Ecoregions

**Figure S2:**
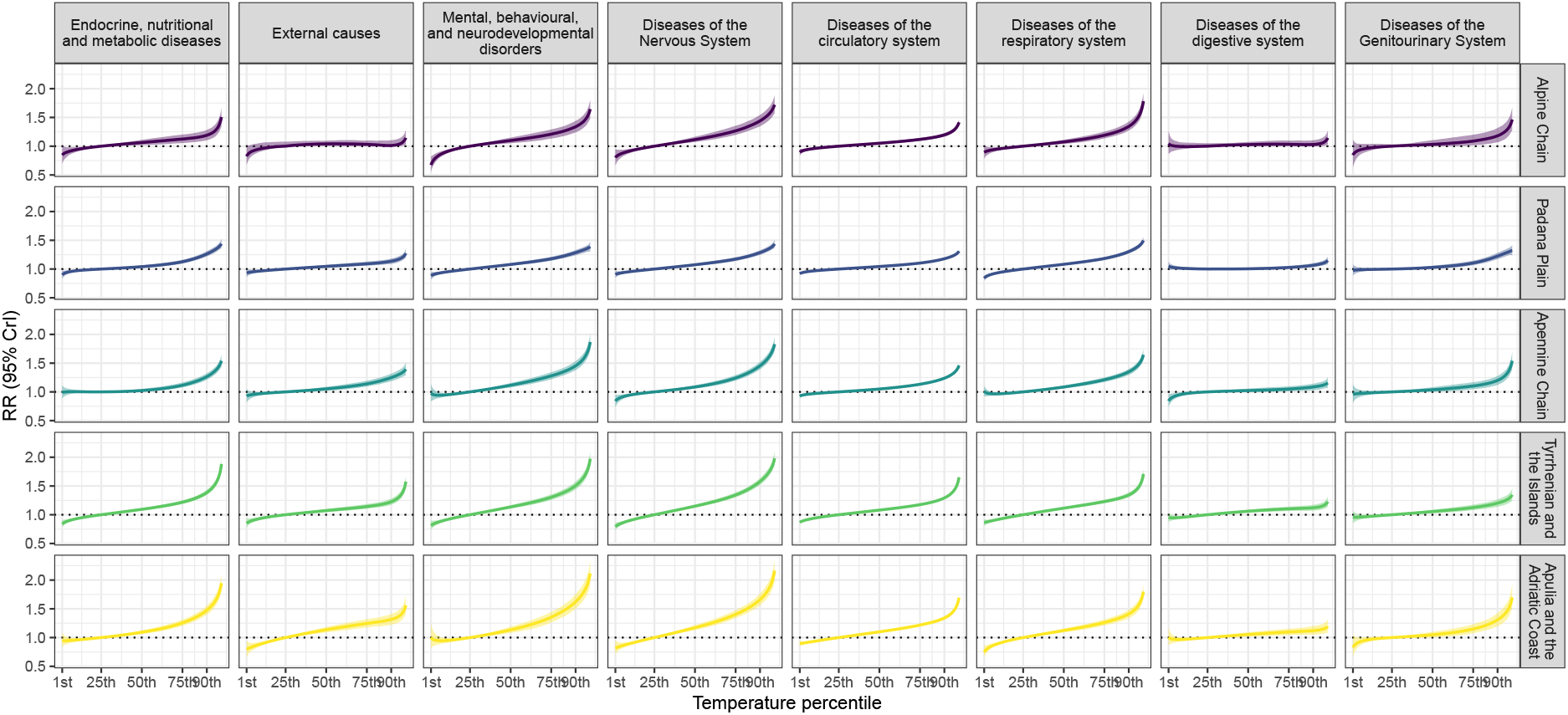
Cause-specific RR estimates for ecoregion-specific daily maximum temperature percentile exposure among individuals aged 65 years and older, by ecoregion. Shaded areas indicate 95% credible intervals. Curves span the 1st–99th percentiles of the temperature distribution.

**Figure S3:**
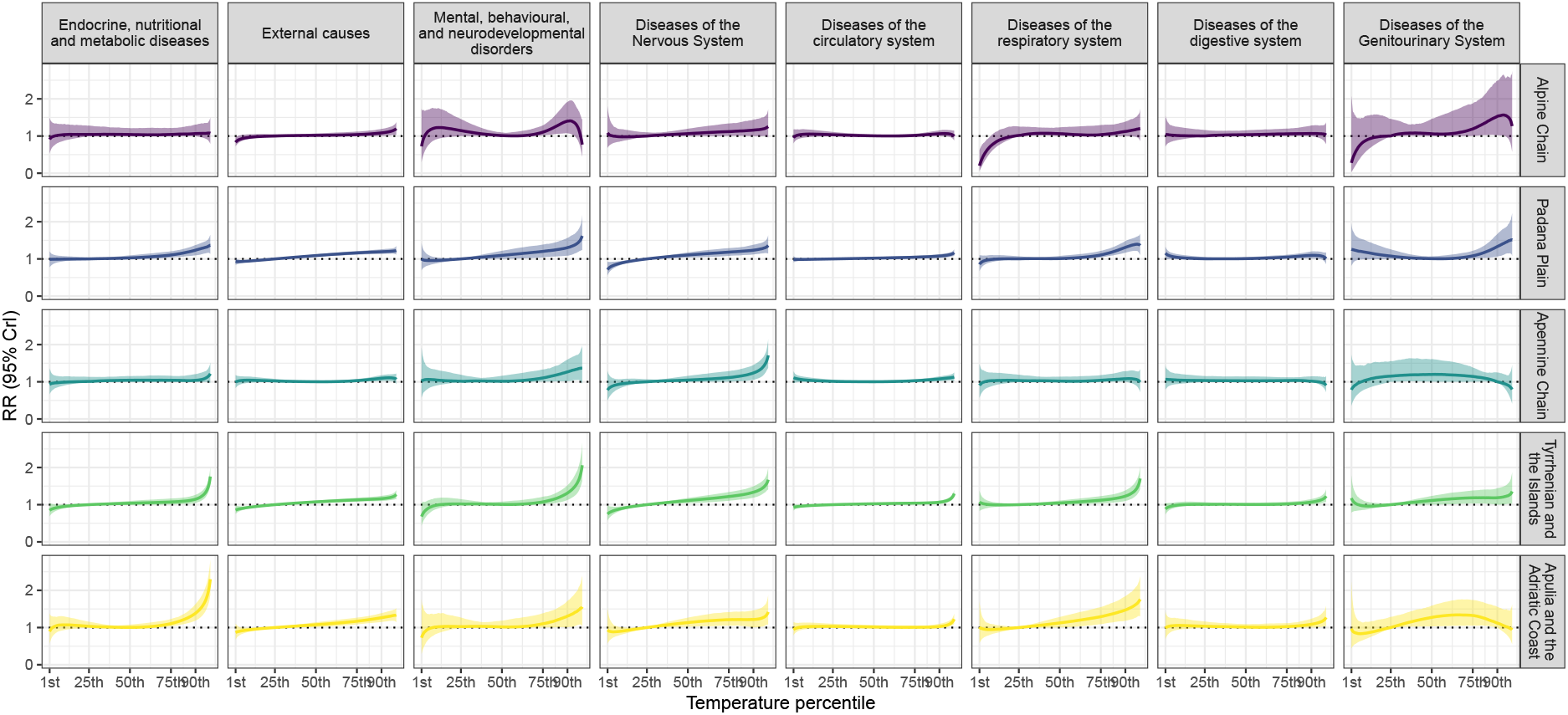
Cause-specific RR estimates for ecoregion-specific daily maximum temperature percentile exposure among individuals aged 18–64 years, by ecoregion. Shaded areas indicate 95% credible intervals. Curves span the 1st–99th percentiles of the temperature distribution.

**Figure S4:**
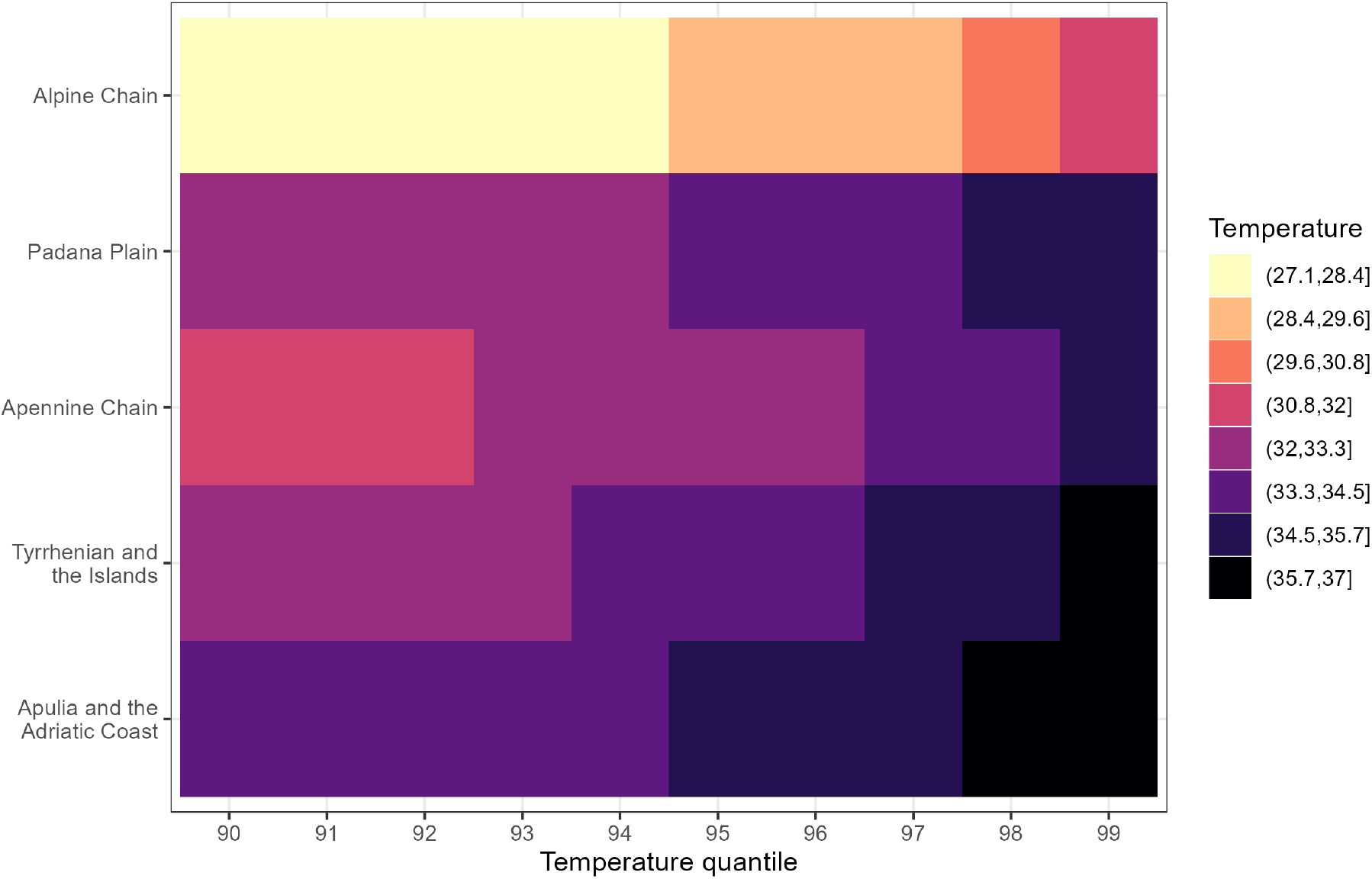
Temperatures corresponding to the ecoregion-specific upper temperature quantiles distribution.

**Figure S5:**
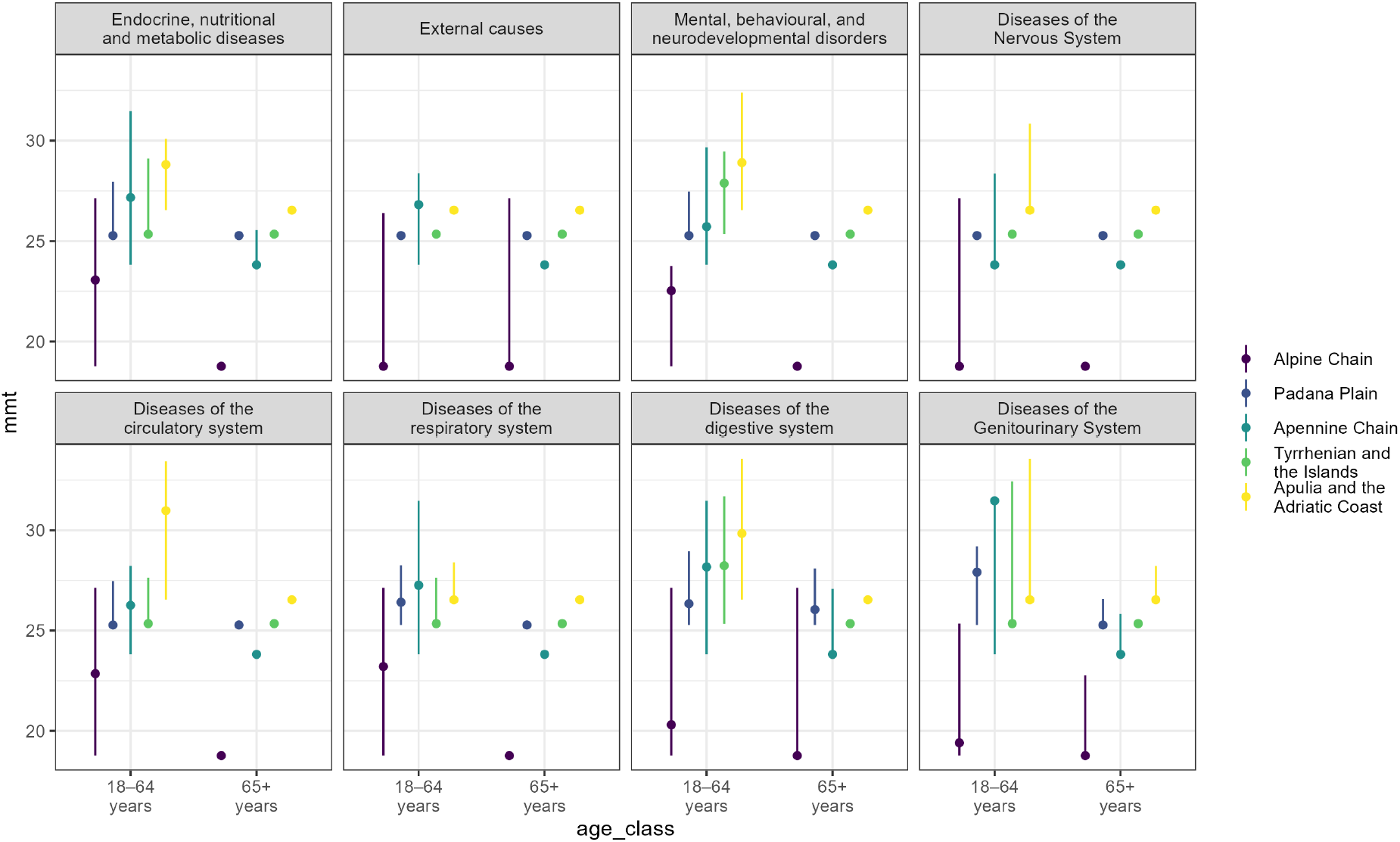
Posterior median estimates of the minimum mortality temperature (MMT) and corresponding 95% credible intervals by ecoregion, age class, and cause of death.

**Figure S6:**
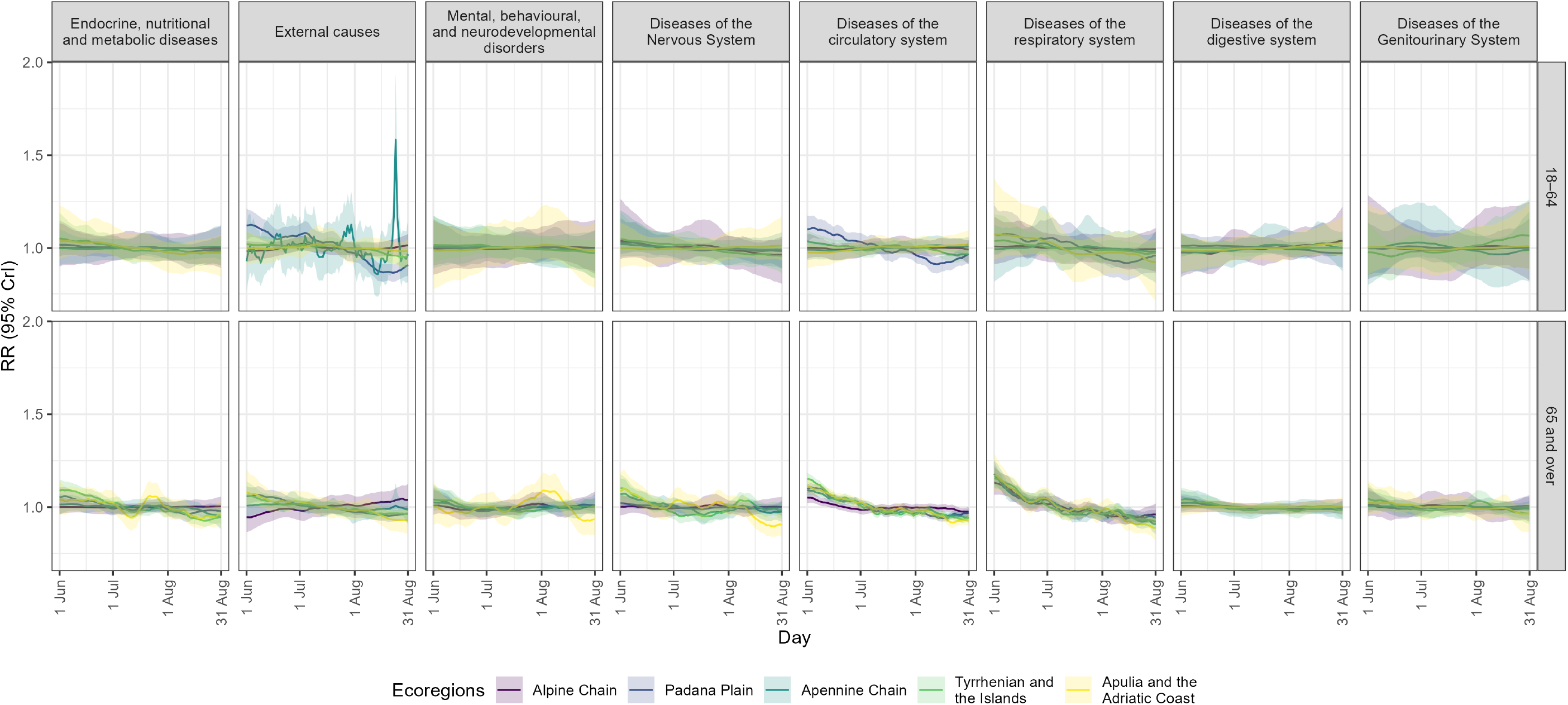
Seasonality trend per ecoregion, cause and age class.

**Figure S7:**
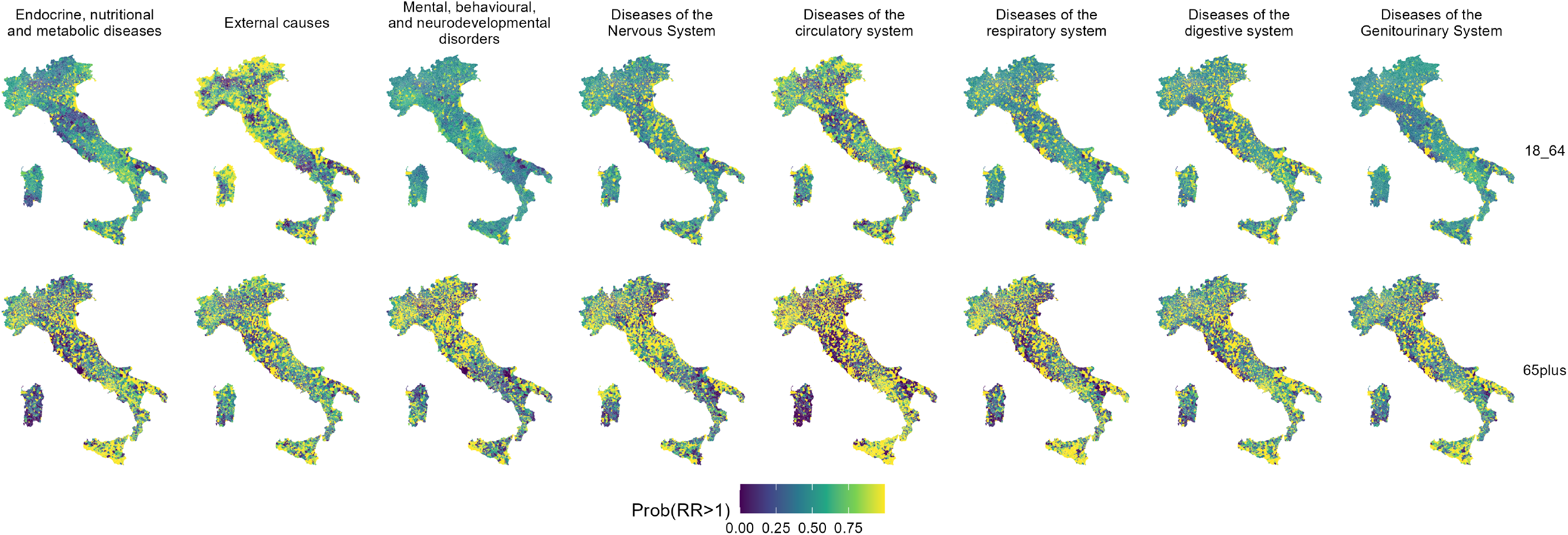
Posterior probability that the RR of the BYM2 component is greater than 1.

**Figure S8:**
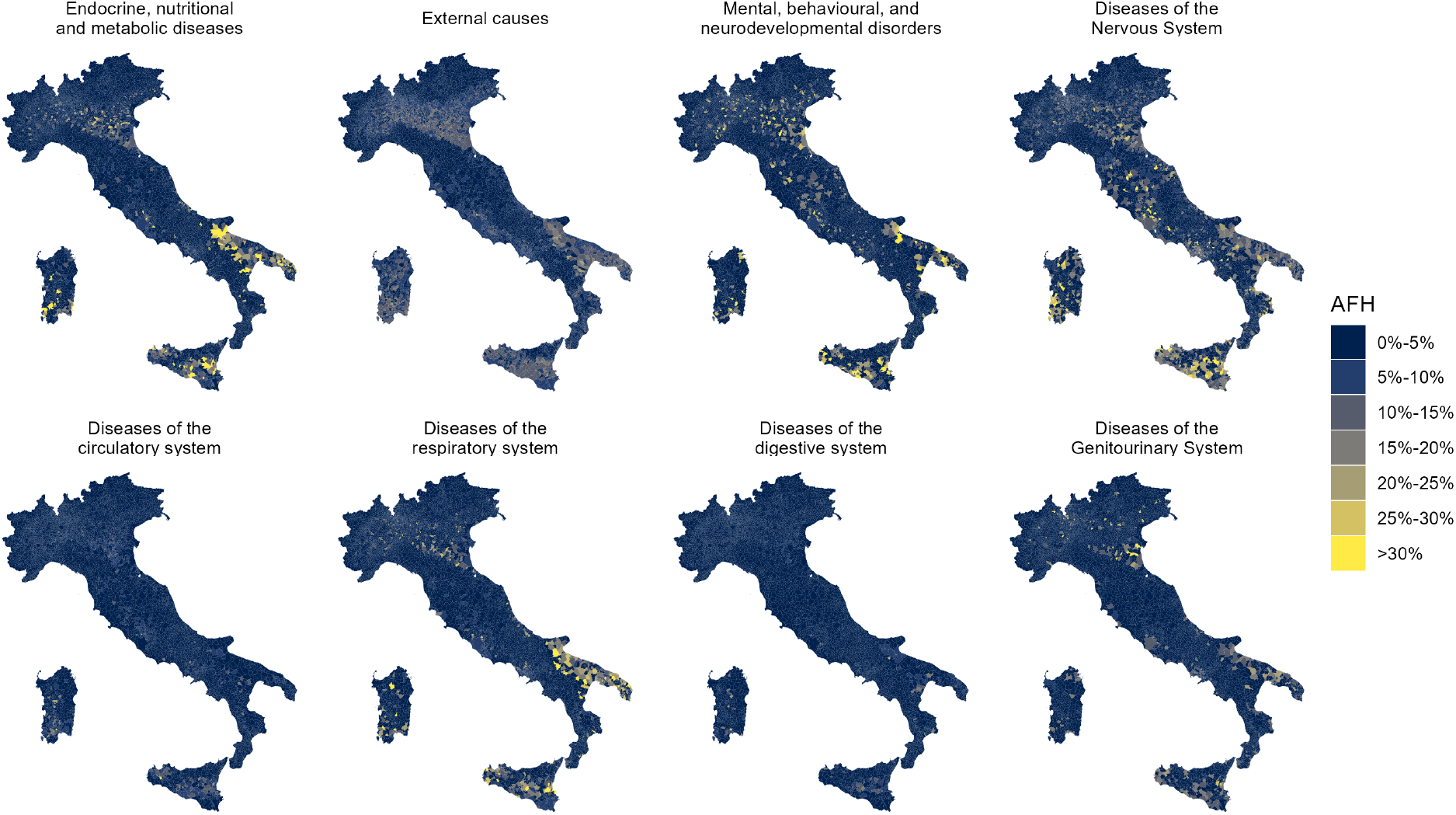
Attributable fraction to heat (AFH) by cause of death among individuals aged 18–64 years, by municipalities. Value represents the posterior median.

**Figure S9:**
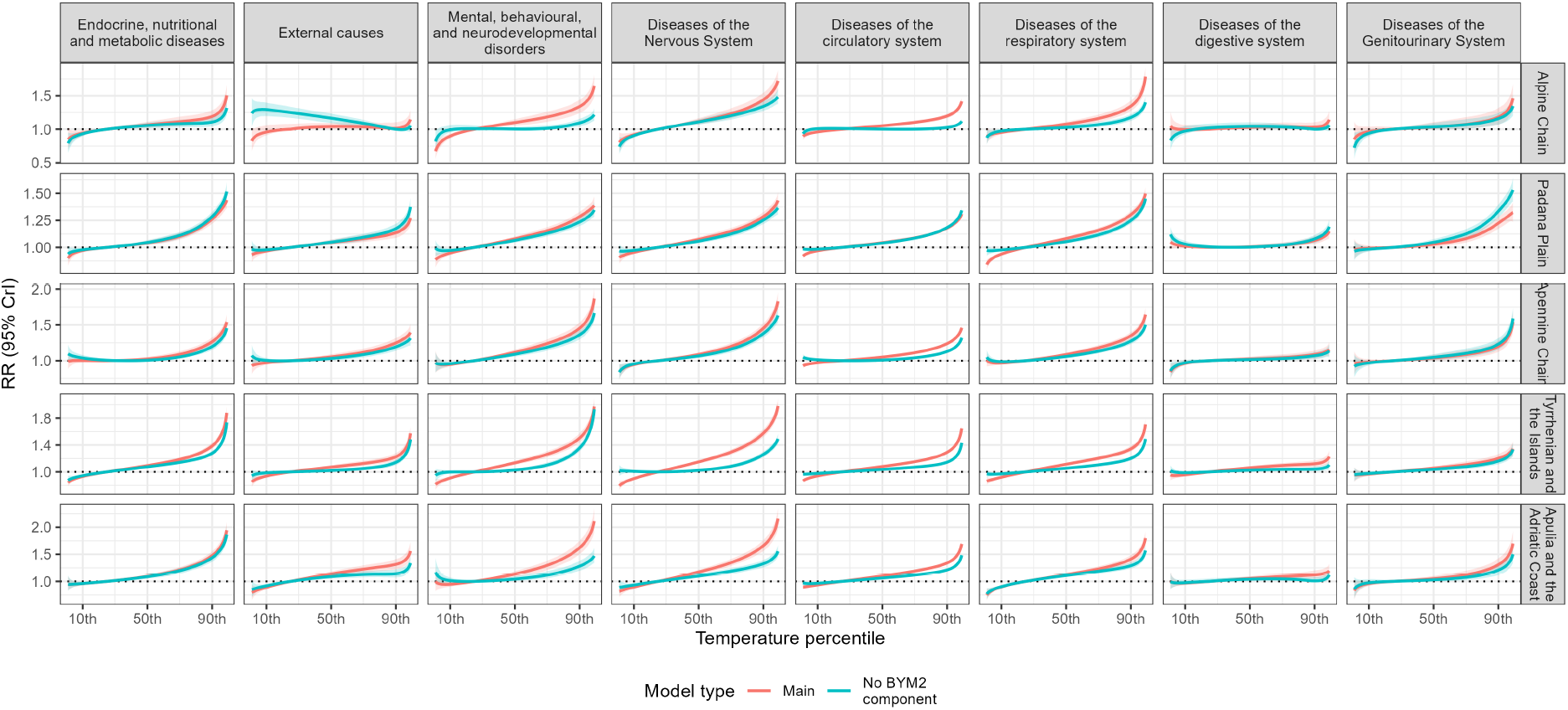
Comparison between the estimated RR in the main model and in the sensitivity model without the BYM2 component.

**Figure S10:**
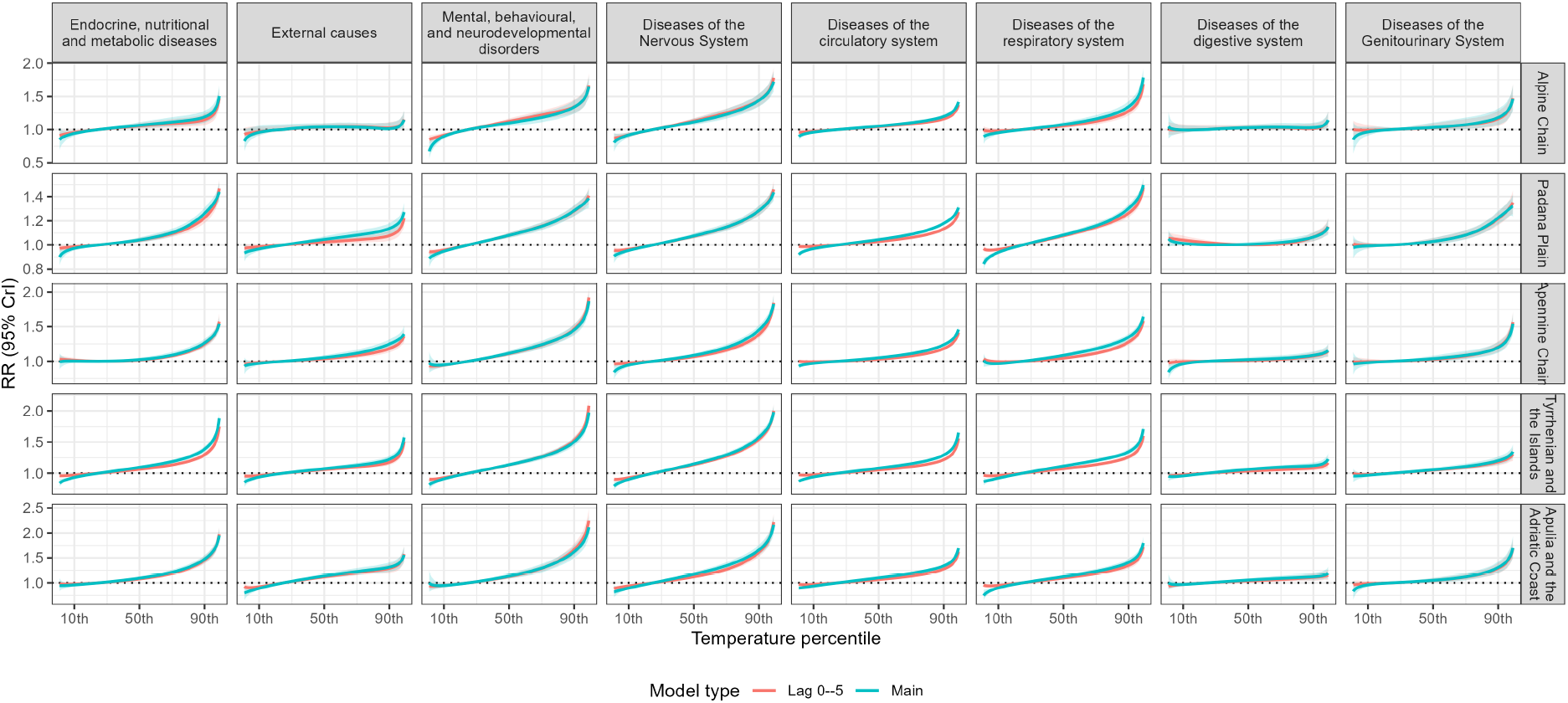
Comparison between the estimated RR in the main model and in the sensitivity model with the exposed window 0–5 days.

**Figure S11:**
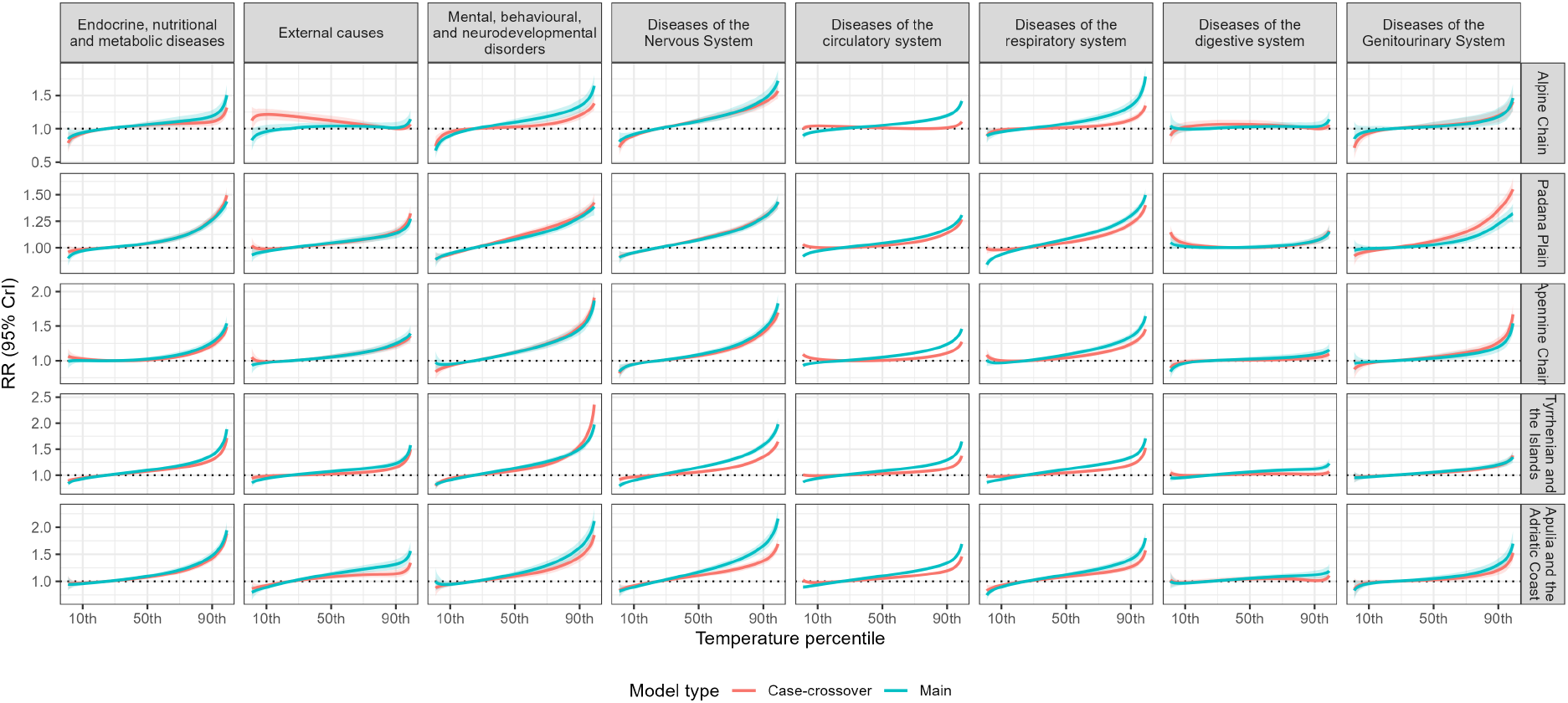
Comparison between the estimated RR in the main model and in the sensitivity model with the case-crossover like design.

**Figure S12:**
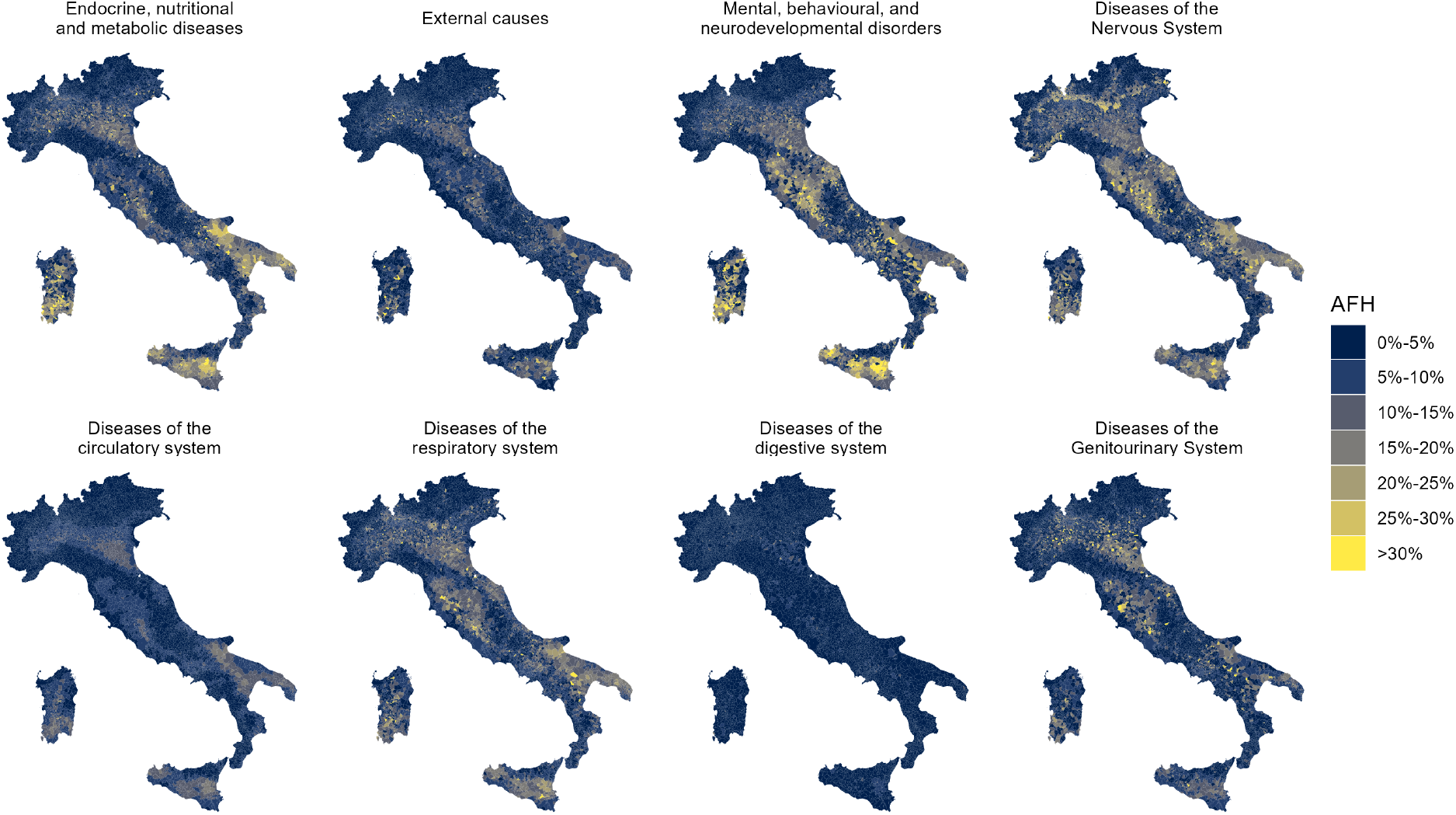
Sensitivity model 1: no BYM2 component. Attributable fraction to heat (AFH) by cause of death among individuals aged 65 years and older, by municipalities. Value represents the posterior median.

**Figure S13:**
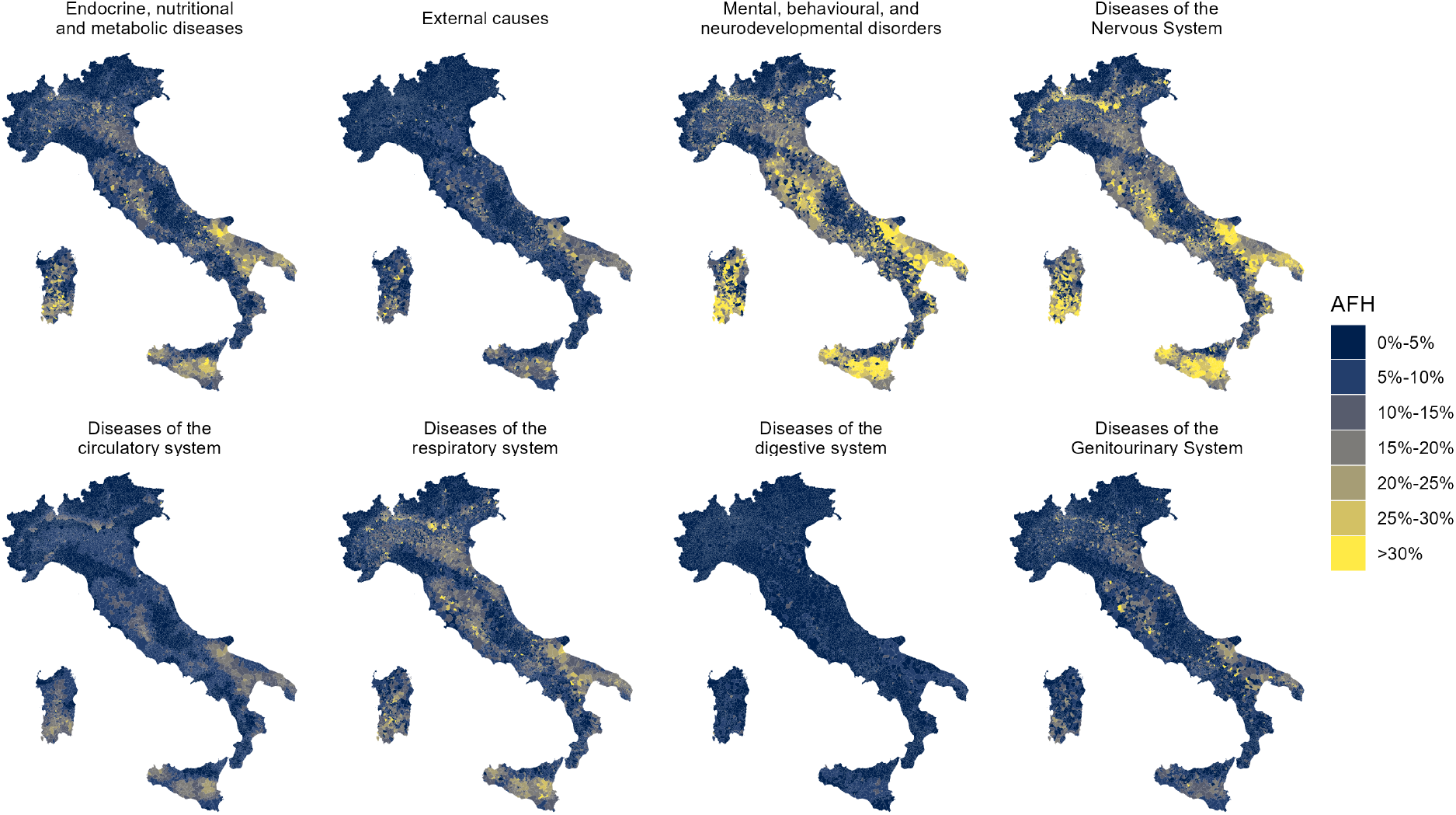
Sensitivity model 2: lag 0–5. Attributable fraction to heat (AFH) by cause of death among individuals aged 65 years and older, by municipalities. Value represents the posterior median.

**Figure S14:**
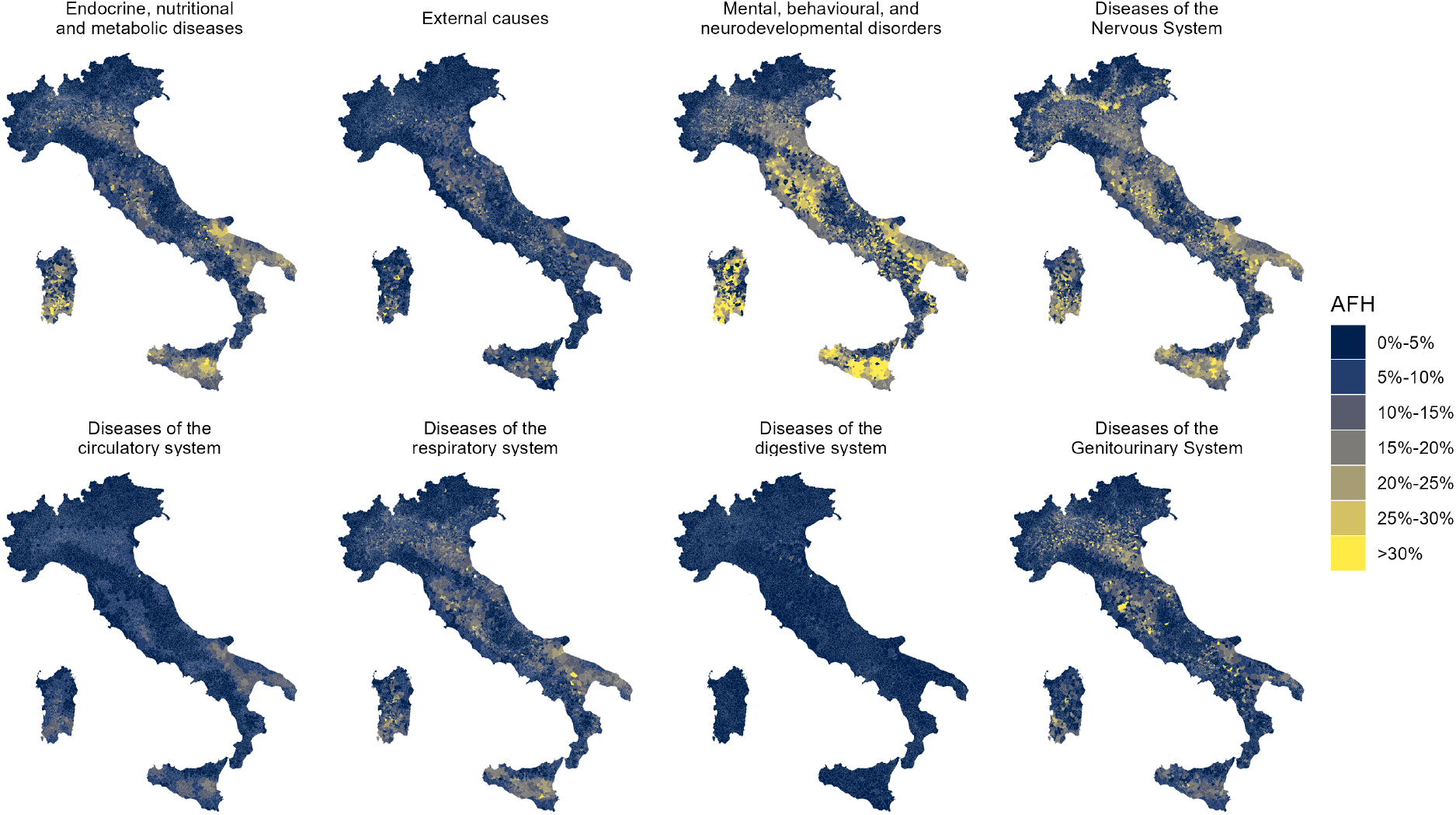
Sensitivity model 3: case-crossover design. Attributable fraction to heat (AFH) by cause of death among individuals aged 65 years and older, by municipalities. Value represents the posterior median.

### S4 Supplementary tables

**Table S1:** Average yearly population, population density, and number of municipalities by ecoregion.

| Ecoregion | 18–64 years | ≥65 years | Population density (km<sup>2</sup>) | No. of municipalities |
| --- | --- | --- | --- | --- |
| Alpine Chain | 3,524,406 | 1,271,483 | 88·2 | 1,846 |
| Padana Plain | 11,867,499 | 4,300,892 | 320·5 | 2,137 |
| Apennine Chain | 5,251,793 | 2,053,149 | 90·8 | 1,686 |
| Tyrrhenian and<br>the Islands | 12,625,566 | 4,068,709 | 194·2 | 1,731 |
| Apulia and the<br>Adriatic Coast | 3,405,093 | 1,133,520 | 150·5 | 464 |
| **Overall Italy** | **36,674,357** | **12,827,753** | **164·3** | **7,864** |

**Table S2:**
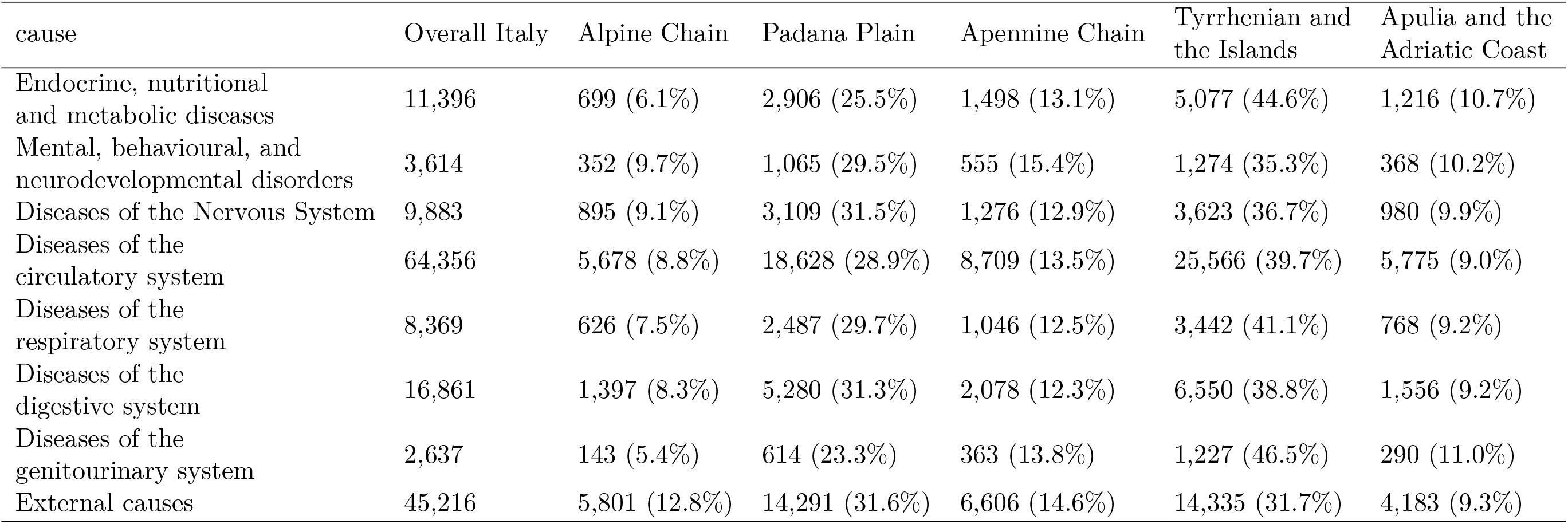
Number of deaths and percentage distribution across ecoregions by cause among individuals aged 18–64 years, 2004–2023.

**Table S3:**
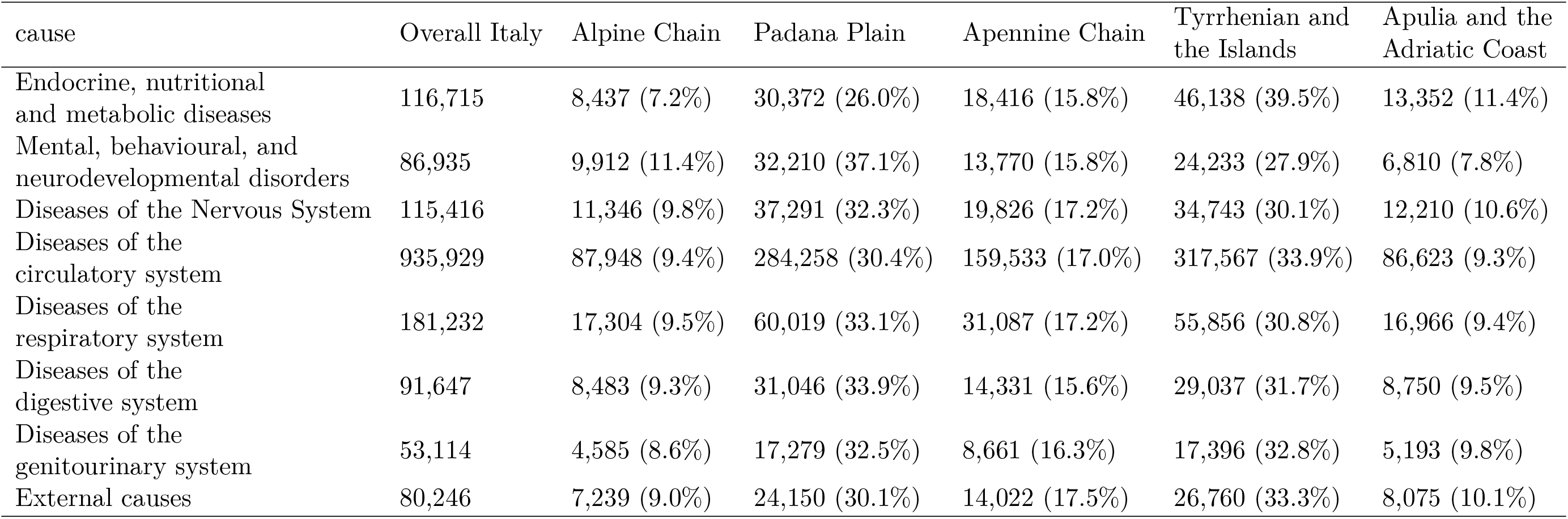
Number of deaths and percentage distribution across ecoregions by cause among individuals aged 65 years or over, 2004–2023.

#### S4.1 Epidemiological metrics

**Table S4:** Attributable fraction (AFH, %) by cause of death and ecoregion among individuals aged 65 years and older. Estimates are posterior medians (95% credible intervals).

| Death cause | Padana Plain | Alpine Chain | Apennine Chain | Tyrrhenian and the Islands | Apulia and the Adriatic Coast | Italy |
| --- | --- | --- | --- | --- | --- | --- |
| Diseases of the circulatory system | 6.40 (5.93 – 6.88) | 8.81 (7.67 – 10.00) | 8.92 (8.18 – 9.62) | 10.24 (9.70 – 10.72) | 12.70 (11.80 – 13.58) | 8.93 (8.64 – 9.21) |
| Diseases of the respiratory system | 10.64 (9.63 – 11.62) | 14.27 (11.46 – 17.00) | 13.18 (11.53 – 14.78) | 12.85 (11.63 – 14.08) | 15.09 (12.96 – 16.95) | 12.50 (11.85 – 13.16) |
| Endocrine, nutritional and metabolic diseases | 8.26 (6.78 – 9.77) | 10.00 (6.00 – 13.39) | 8.93 (6.97 – 11.04) | 13.08 (11.78 – 14.33) | 15.78 (13.35 – 17.86) | 11.25 (10.44 – 12.11) |
| Mental and behavioural disorders | 10.24 (8.76 – 11.71) | 14.64 (11.12 – 18.17) | 17.43 (15.09 – 19.75) | 17.49 (15.81 – 19.19) | 19.35 (16.36 – 22.27) | 14.63 (13.61 – 15.64) |
| Diseases of the genitourinary system | 7.00 (5.13 – 9.02) | 8.28 (3.49 – 13.51) | 8.54 (5.38 – 11.78) | 7.53 (5.07 – 9.65) | 10.19 (6.59 – 13.87) | 7.85 (6.61 – 9.10) |
| Diseases of the nervous system | 10.11 (8.82 – 11.33) | 18.22 (14.89 – 21.65) | 15.81 (13.77 – 17.84) | 17.71 (16.33 – 19.09) | 20.58 (18.35 – 22.57) | 15.27 (14.48 – 16.07) |
| Diseases of the digestive system | 1.95 (1.02 – 2.92) | 2.48 (0.22 – 6.53) | 3.65 (1.25 – 6.26) | 5.81 (4.04 – 7.45) | 5.69 (2.98 – 8.20) | 3.86 (2.90 – 4.74) |
| External causes of mortality | 5.81 (4.09 – 7.40) | 2.51 (0.21 – 6.79) | 9.21 (6.88 – 11.51) | 9.04 (7.28 – 10.76) | 13.33 (10.45 – 16.13) | 7.94 (7.06 – 8.92) |

**Table S5:** Heat-attributable deaths (ECH) by cause of death and ecoregion among individuals aged 65 years and older. Estimates are posterior medians (95% credible intervals).

| Death cause | Padana Plain | Alpine Chain | Apennine Chain | Tyrrhenian and the Islands | Apulia and the Adriatic Coast | Italy |
| --- | --- | --- | --- | --- | --- | --- |
| Diseases of the circulatory system | 18191 (16869 – 19552) | 7746 (6746 – 8796) | 14231 (13046 – 15354) | 32505 (30802 – 34053) | 10998 (10223 – 11760) | 83624 (80819 – 86224) |
| Diseases of the respiratory system | 6384 (5779 – 6973) | 2469 (1983 – 2942) | 4096 (3584 – 4595) | 7177 (6493 – 7864) | 2559 (2198 – 2876) | 22654 (21479 – 23846) |
| Endocrine, nutritional and metabolic diseases | 2509 (2060 – 2967) | 843 (506 – 1130) | 1645 (1284 – 2032) | 6035 (5435 – 6614) | 2108 (1783 – 2384) | 13131 (12182 – 14138) |
| Mental and behavioural disorders | 3298 (2821 – 3771) | 1451 (1102 – 1801) | 2399 (2077 – 2720) | 4238 (3831 – 4649) | 1318 (1114 – 1517) | 12720 (11831 – 13593) |
| Diseases of the genitourinary system | 1210 (886 – 1559) | 380 (160 – 620) | 740 (466 – 1020) | 1311 (882 – 1679) | 529 (342 – 720) | 4171 (3511 – 4836) |
| Diseases of the nervous system | 3769 (3289 – 4224) | 2067 (1689 – 2457) | 3134 (2729 – 3536) | 6153 (5674 – 6631) | 2513 (2240 – 2756) | 17622 (16708 – 18549) |
| Diseases of the digestive system | 604 (315 – 906) | 211 (19 – 554) | 523 (179 – 898) | 1686 (1174 – 2163) | 498 (261 – 718) | 3537 (2656 – 4344) |
| External causes of mortality | 1402 (988 – 1786) | 182 (15 – 492) | 1291 (964 – 1614) | 2419 (1948 – 2881) | 1077 (844 – 1302) | 6373 (5663 – 7159) |

**Table S6:** Excess deaths per 1,000 inhabitants (ERH) by cause of death and ecoregion among individuals aged 65 years and older. Estimates are posterior medians (95% credible intervals).

| Death cause | Padana Plain | Alpine Chain | Apennine Chain | Tyrrhenian and the Islands | Apulia and the Adriatic Coast | Italy |
| --- | --- | --- | --- | --- | --- | --- |
| Diseases of the circulatory system | 4.23 (3.92 – 4.55) | 6.09 (5.31 – 6.92) | 6.93 (6.35 – 7.48) | 7.99 (7.57 – 8.37) | 9.70 (9.02 – 10.37) | 6.52 (6.30 – 6.72) |
| Diseases of the respiratory system | 1.48 (1.34 – 1.62) | 1.94 (1.56 – 2.31) | 2.00 (1.75 – 2.24) | 1.76 (1.60 – 1.93) | 2.26 (1.94 – 2.54) | 1.77 (1.67 – 1.86) |
| Endocrine, nutritional and metabolic diseases | 0.58 (0.48 – 0.69) | 0.66 (0.40 – 0.89) | 0.80 (0.63 – 0.99) | 1.48 (1.34 – 1.63) | 1.86 (1.57 – 2.10) | 1.02 (0.95 – 1.10) |
| Mental and behavioural disorders | 0.77 (0.66 – 0.88) | 1.14 (0.87 – 1.42) | 1.17 (1.01 – 1.32) | 1.04 (0.94 – 1.14) | 1.16 (0.98 – 1.34) | 0.99 (0.92 – 1.06) |
| Diseases of the genitourinary system | 0.28 (0.21 – 0.36) | 0.30 (0.13 – 0.49) | 0.36 (0.23 – 0.50) | 0.32 (0.22 – 0.41) | 0.47 (0.30 – 0.64) | 0.33 (0.27 – 0.38) |
| Diseases of the nervous system | 0.88 (0.76 – 0.98) | 1.63 (1.33 – 1.93) | 1.53 (1.33 – 1.72) | 1.51 (1.39 – 1.63) | 2.22 (1.98 – 2.43) | 1.37 (1.30 – 1.45) |
| Diseases of the digestive system | 0.14 (0.07 – 0.21) | 0.17 (0.02 – 0.44) | 0.25 (0.09 – 0.44) | 0.41 (0.29 – 0.53) | 0.44 (0.23 – 0.63) | 0.28 (0.21 – 0.34) |
| External causes of mortality | 0.33 (0.23 – 0.42) | 0.14 (0.01 – 0.39) | 0.63 (0.47 – 0.79) | 0.59 (0.48 – 0.71) | 0.95 (0.74 – 1.15) | 0.50 (0.44 – 0.56) |

**Table S7:** Attributable fraction (AFH, %) by cause of death and ecoregion among individuals aged 18–64 years. Estimates are posterior medians (95% credible intervals).

| Death cause | Padana Plain | Alpine Chain | Apennine Chain | Tyrrhenian and the Islands | Apulia and the Adriatic Coast | Italy |
| --- | --- | --- | --- | --- | --- | --- |
| Diseases of the circulatory system | 2.98 (1.32 – 4.83) | 1.07 (0.00 – 2.41) | 2.20 (0.64 – 3.96) | 3.33 (1.39 – 5.34) | 1.31 (0.50 – 2.14) | 2.71 (1.75 – 3.69) |
| Diseases of the respiratory system | 6.69 (3.40 – 11.41) | 4.12 (0.01 – 16.60) | 1.59 (0.00 – 5.83) | 8.05 (3.81 – 12.65) | 15.82 (6.55 – 24.12) | 7.40 (5.04 – 10.13) |
| Endocrine, nutritional and metabolic diseases | 6.70 (3.09 – 10.84) | 0.90 (0.00 – 3.52) | 2.49 (0.05 – 9.18) | 6.71 (2.93 – 10.69) | 11.08 (7.19 – 14.64) | 6.41 (4.37 – 8.61) |
| Mental and behavioural disorders | 11.40 (4.32 – 18.50) | 6.57 (1.30 – 11.59) | 7.26 (1.00 – 13.30) | 7.40 (3.69 – 14.16) | 6.77 (0.48 – 17.43) | 8.59 (5.72 – 12.03) |
| Diseases of the genitourinary system | 6.79 (1.79 – 11.20) | 13.27 (0.11 – 34.75) | 0.00 (0.00 – 0.82) | 9.89 (2.20 – 17.67) | 13.51 (1.36 – 24.95) | 8.37 (4.41 – 12.47) |
| Diseases of the nervous system | 10.15 (5.68 – 14.10) | 7.93 (0.24 – 16.59) | 9.99 (3.39 – 17.15) | 12.67 (7.83 – 17.13) | 11.41 (3.86 – 18.28) | 10.90 (8.31 – 13.64) |
| Diseases of the digestive system | 2.08 (0.36 – 3.99) | 2.32 (0.00 – 9.84) | 0.00 (0.00 – 0.44) | 1.58 (0.20 – 4.82) | 2.44 (0.32 – 5.46) | 1.82 (0.83 – 3.25) |
| External causes of mortality | 8.22 (6.25 – 10.24) | 2.65 (0.28 – 6.55) | 2.22 (0.49 – 3.93) | 7.54 (5.21 – 10.04) | 9.73 (5.80 – 13.37) | 6.57 (5.52 – 7.67) |

**Table S8:**
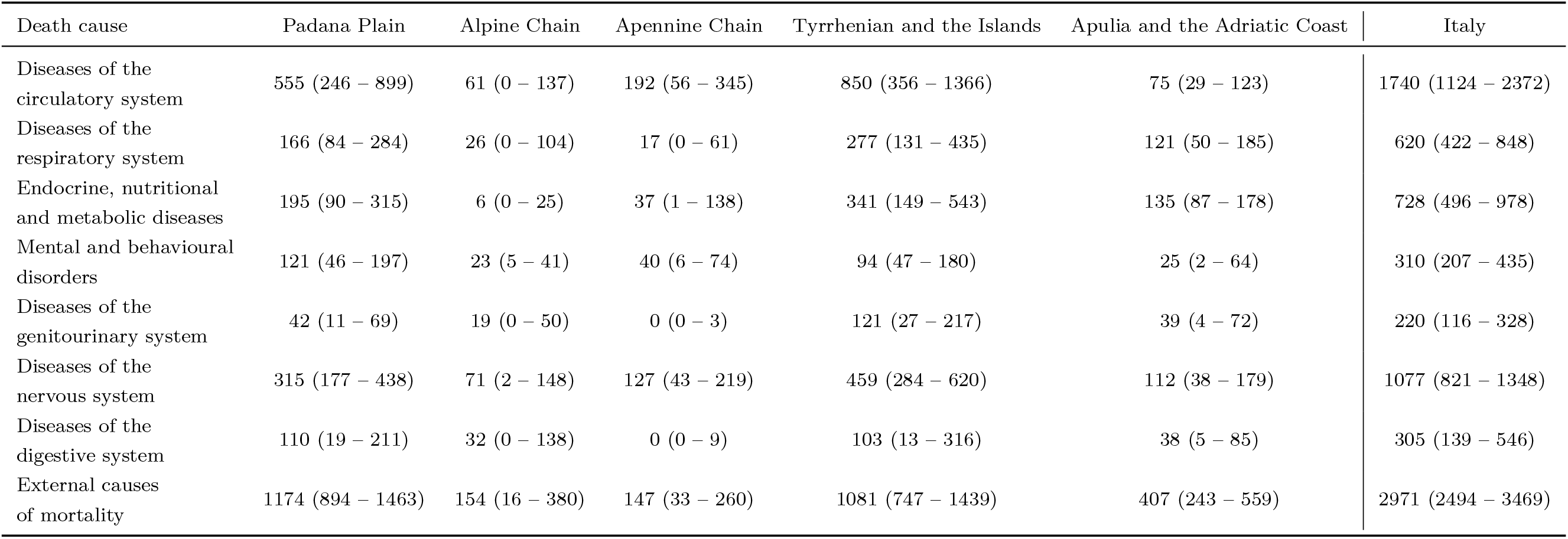
Heat-attributable deaths (ECH) by cause of death and ecoregion among individuals aged 18–64 years. Estimates are posterior medians (95% credible intervals).

| Death cause | Padana Plain | Alpine Chain | Apennine Chain | Tyrrhenian and the Islands | Apulia and the Adriatic Coast | Italy |
| --- | --- | --- | --- | --- | --- | --- |
| Diseases of the circulatory system | 555 (246 – 899) | 61 (0 – 137) | 192 (56 – 345) | 850 (356 – 1366) | 75 (29 – 123) | 1740 (1124 – 2372) |
| Diseases of the respiratory system | 166 (84 – 284) | 26 (0 – 104) | 17 (0 – 61) | 277 (131 – 435) | 121 (50 – 185) | 620 (422 – 848) |
| Endocrine, nutritional and metabolic diseases | 195 (90 – 315) | 6 (0 – 25) | 37 (1 – 138) | 341 (149 – 543) | 135 (87 – 178) | 728 (496 – 978) |
| Mental and behavioural disorders | 121 (46 – 197) | 23 (5 – 41) | 40 (6 – 74) | 94 (47 – 180) | 25 (2 – 64) | 310 (207 – 435) |
| Diseases of the genitourinary system | 42 (11 – 69) | 19 (0 – 50) | 0 (0 – 3) | 121 (27 – 217) | 39 (4 – 72) | 220 (116 – 328) |
| Diseases of the nervous system | 315 (177 – 438) | 71 (2 – 148) | 127 (43 – 219) | 459 (284 – 620) | 112 (38 – 179) | 1077 (821 – 1348) |
| Diseases of the digestive system | 110 (19 – 211) | 32 (0 – 138) | 0 (0 – 9) | 103 (13 – 316) | 38 (5 – 85) | 305 (139 – 546) |
| External causes of mortality | 1174 (894 – 1463) | 154 (16 – 380) | 147 (33 – 260) | 1081 (747 – 1439) | 407 (243 – 559) | 2971 (2494 – 3469) |

**Table S9:** Excess deaths per 1,000 inhabitants (ERH) by cause of death and ecoregion among individuals aged 18–64 years. Estimates are posterior medians (95% credible intervals).

| Death cause | Padana Plain | Alpine Chain | Apennine Chain | Tyrrhenian and the Islands | Apulia and the Adriatic Coast | Italy |
| --- | --- | --- | --- | --- | --- | --- |
| Diseases of the circulatory system | 0.05 (0.02 – 0.08) | 0.02 (0.00 – 0.04) | 0.04 (0.01 – 0.07) | 0.07 (0.03 – 0.11) | 0.02 (0.01 – 0.04) | 0.05 (0.03 – 0.06) |
| Diseases of the respiratory system | 0.01 (0.01 – 0.02) | 0.01 (0.00 – 0.03) | 0.00 (0.00 – 0.01) | 0.02 (0.01 – 0.03) | 0.04 (0.01 – 0.05) | 0.02 (0.01 – 0.02) |
| Endocrine, nutritional and metabolic diseases | 0.02 (0.01 – 0.03) | 0.00 (0.00 – 0.01) | 0.01 (0.00 – 0.03) | 0.03 (0.01 – 0.04) | 0.04 (0.03 – 0.05) | 0.02 (0.01 – 0.03) |
| Mental and behavioural disorders | 0.01 (0.00 – 0.02) | 0.01 (0.00 – 0.01) | 0.01 (0.00 – 0.01) | 0.01 (0.00 – 0.01) | 0.01 (0.00 – 0.02) | 0.01 (0.01 – 0.01) |
| Diseases of the genitourinary system | 0.00 (0.00 – 0.01) | 0.01 (0.00 – 0.01) | 0.00 (0.00 – 0.00) | 0.01 (0.00 – 0.02) | 0.01 (0.00 – 0.02) | 0.01 (0.00 – 0.01) |
| Diseases of the nervous system | 0.03 (0.01 – 0.04) | 0.02 (0.00 – 0.04) | 0.02 (0.01 – 0.04) | 0.04 (0.02 – 0.05) | 0.03 (0.01 – 0.05) | 0.03 (0.02 – 0.04) |
| Diseases of the digestive system | 0.01 (0.00 – 0.02) | 0.01 (0.00 – 0.04) | 0.00 (0.00 – 0.00) | 0.01 (0.00 – 0.02) | 0.01 (0.00 – 0.02) | 0.01 (0.00 – 0.01) |
| External causes of mortality | 0.10 (0.08 – 0.12) | 0.04 (0.00 – 0.11) | 0.03 (0.01 – 0.05) | 0.09 (0.06 – 0.11) | 0.12 (0.07 – 0.16) | 0.08 (0.07 – 0.09) |

**Table S10:** Difference in WAIC between the main model and the sensitivity model without the BYM2 component among individuals aged 65 years or older. Negative values indicate a lower WAIC for the main model.

| Death cause | Padana Plain | Alpine Chain | Apennine Chain | Tyrrhenian and<br>the Islands | Apulia and the<br>Adriatic Coast |
| --- | --- | --- | --- | --- | --- |
| Diseases of the<br>circulatory system | -108504 | -41446 | -39949 | -44390 | -13186 |
| Diseases of the<br>respiratory system | -42034 | -16202 | -16066 | -14382 | -4578 |
| Endocrine, nutritional<br>and metabolic diseases | -8708 | -3030 | -4700 | -6903 | -1768 |
| Mental and behavioural<br>disorders | -17723 | -7008 | -4959 | -5913 | -1386 |
| Diseases of the<br>genitourinary system | -11771 | -3907 | -5087 | -4292 | -1152 |
| Diseases of the<br>nervous system | -12700 | -4942 | -4641 | -5840 | -2123 |
| Diseases of the<br>digestive system | -30908 | -11104 | -13187 | -13034 | -4623 |
| External causes<br>of mortality | -10829 | -3904 | -365235 | -7235 | -2204 |

**Table S11:** Difference in WAIC between the main model and the sensitivity model with the case-crossover design among individuals aged 65 years or older. Negative values indicate a lower WAIC for the main model.

| Death cause | Padana Plain | Alpine Chain | Apennine Chain | Tyrrhenian and<br>the Islands | Apulia and the<br>Adriatic Coast |
| --- | --- | --- | --- | --- | --- |
| Diseases of the<br>circulatory system | -82334 | -33966 | -32645 | -38061 | -10944 |
| Diseases of the<br>respiratory system | -38205 | -14674 | -14386 | -14118 | -4215 |
| Endocrine, nutritional<br>and metabolic diseases | -8555 | -2926 | -4598 | -6222 | -1740 |
| Mental and behavioural<br>disorders | -18013 | -7211 | -5880 | -7394 | -1928 |
| Diseases of the<br>genitourinary system | -11879 | -3764 | -5063 | -4300 | -1126 |
| Diseases of the<br>nervous system | -13048 | -4830 | -4678 | -5961 | -2149 |
| Diseases of the<br>digestive system | -29278 | -10268 | -12299 | -12915 | -4368 |
| External causes<br>of mortality | -10398 | -3735 | -1757 | -7073 | -2084 |

## Notes

### Competing Interest Statement

The authors have declared no competing interest.

